# Population-scale detection of methylation outliers from long-read genome sequencing

**DOI:** 10.64898/2026.06.09.26355279

**Authors:** Tanner D Jensen, Rhina Kaur, Devon E Bonner, Jon Nguyen, Chloe M Reuter, Undiagnosed Diseases Network, Genomics Research to Elucidate the Genetics of Rare Diseases (GREGoR) Consortium, Euan A. Ashley, Jonathan A. Bernstein, Matthew T. Wheeler, Stephen B Montgomery

## Abstract

**Background:** Aberrant DNA methylation can mediate the functional effects of rare genetic variation and contribute to imprinting disorders, repeat expansion diseases, and other pathogenic regulatory mechanisms. Long-read sequencing technologies now enable genome-wide detection of CpG methylation alongside genetic variation from a single assay. However, methods for systematic identification and interpretation of methylation outliers from long-read sequencing data remain limited.

**Methods:** We developed METAFORA, a computational workflow for detecting methylation outlier regions from PacBio and Oxford Nanopore long-read sequencing data. METAFORA constructs population-level methylation references, segments the genome into correlated CpG blocks, infers technical and biological sources of variation through hidden factor estimation, models uncertainty due to variable depth sequencing, and computes covariate-adjusted methylation outlier scores for individual samples. We applied METAFORA across large long-read sequencing cohorts and integrated methylation outliers with multi-omic data. METAFORA is implemented as a snakemake workflow available at https://github.com/tjense25/METAFORA.

**Results:** METAFORA identified methylation outlier regions associated with rare structural variants, tandem repeat expansions, and imprinting abnormalities. We found outlier regions were enriched for molecular outliers across transcriptomic and chromatin accessibility datasets, supporting their functional relevance in gene regulation. In a representative case, METAFORA identified an imprinting defect affecting the GNAS locus associated with an STX16 deletion.

**Conclusions:** METAFORA enables scalable detection and interpretation of methylation outliers from long-read sequencing data and provides a framework for integrating epigenetic outliers with genomic and multi-omic analyses. These approaches may improve interpretation of rare regulatory variation and support discovery of clinically relevant epigenetic abnormalities in genomic medicine.

## Background

Despite advances in genome sequencing, many rare disease cases remain undiagnosed because functional effects of variants are difficult to interpret (1). Molecular outlier analysis—detecting individuals with genes whose molecular trait levels deviate substantially from the population norm—has emerged as a powerful strategy for revealing dysregulated genes and linking them to rare variants. Outlier detection from RNA-seq data—both for expression and splicing—has been applied in rare disease contexts (2–5), and similar strategies have been extended to proteomics and chromatin accessibility (ATAC-seq) data (6–8).

Methylation outlier analysis offers an additional, complementary layer for assessing variant effects. Further, beyond rare variant effects, methylation outlier detection is valuable for diagnosing imprinting disorders, which involve aberrant methylation at loci where one allele is normally silenced based on parental origin. Such effects can result in abnormal gene expression and disorders such as Prader Willi or Silver-Russell Syndrome, with a range of features including growth and endocrine abnormalities, neurodevelopmental delay, and distinctive facial morphology (9).

Previous studies and clinical assays that have called methylation outliers rely on methylation arrays, which assay a limited set of CpG sites genome-wide (10). Outliers are called at individual CpGs or aggregated within promoter boundaries, making regulatory interpretation difficult (11,12). Other approaches for methylation dysregulation analyses bin the genome into fixed tiles, which can dilute or fragment signals when outliers span bin boundaries or occupy only a small fraction of a bin (13,14), or have focused solely on CpG islands or other genome annotations (15,16). Array-based profiling has also been used to predict imprinting disorders and define “episignatures” for diverse syndromes such as those linked to chromatin remodeling genes (17–19). These “episignatures” have now demonstrated clinical utility in over 100 specific disorders (17,20,https://episign.com) However, this approach depends on pre-trained supervised models from known case–control datasets, limiting its ability to detect novel patterns in previously uncharacterized rare diseases.

Long-read genome sequencing (lrGS) offers an opportunity to address these limitations. In addition to improved mappability across repetitive regions, multiple lrGS platforms–including PacBio and Oxford Nanopore Technology (ONT)--can detect CpG methylation genome-wide, providing far broader coverage than the ∼80,000 CpGs profiled by arrays. Although error profiles vary by platform and software (21,22), long-read sequencing has clear promise for detecting disease-relevant epigenetic changes (23–25). These initial studies have relied on single individuals or families which do not provide enough samples to call outliers, yet the production of long-read data is rapidly scaling up through initiatives such as GREGoR, GA4K (“Genomics Answers for Kids”), and All of Us (26–29)—creating the need for tools that can jointly analyze methylation data across larger cohorts and from diverse platforms.

Here, we present METAFORA, a framework for detecting methylation outlier regions from population-scale long-read datasets. METAFORA segments the genome into correlated CpG blocks learned directly from the data, avoiding the constraints of fixed bins or predefined annotations. It infers hidden factor covariates capturing biological (e.g. sex, age, cell-type composition) and technical (e.g. platform- or software-specific) variation, enabling robust adjustment and joint analysis across platforms. It also can model uncertainty from variable coverage sequencing, better accounting for long-read error modes. This approach can therefore increase reference sample size, improve statistical power, and yield interpretable, high-resolution maps of local methylation deviations relevant to rare disease discovery. We test METAFORA on both simulated and real data from diverse sources and show its potential to identify biologically-relevant methylation changes in clinical contexts.

## Methods

We developed METAFORA, a statistical framework for detecting methylation outlier regions from lrGS CpG methylation data. METAFORA leverages tissue-matched population references, depth-aware statistical modeling, and covariate adjustment to identify rare, sample-specific methylation deviations while accounting for variability inherent to lrGS.

Aligned lrGS BAM files containing CpG methylation tags are converted into CpG-level methylation and depth profiles. For each tissue, METAFORA constructs a population reference representing expected methylation levels across the cohort. To address the high dimensionality of >36 million CpGs and improve statistical stability, the genome is segmented into correlated CpG blocks using change-point detection, and methylation is summarized at the segment level.

To detect candidate events, METAFORA compares each sample to the tissue reference using depth-aware deviance statistics, identifying contiguous regions of significant deviation via segmentation. Segment-level methylation values are then residualized against inferred hidden factors and user-supplied covariates (e.g., age, sex, batch, sequencing depth) prior to z-score–based outlier calling. Sex chromosomes are analyzed using sex-stratified references, and global outlier samples can be automatically identified and excluded.

Because recurrent events may be represented by slightly different segment boundaries across individuals, METAFORA additionally performs joint outlier calling by harmonizing overlapping segments across samples and recalculating cohort-level statistics.

Identified outliers are functionally annotated using gene models and regulatory genomic tracks, including CpG islands, open chromatin regions, predicted Activity-by-Contact enhancers, and curated imprinting disorder differently methylated regions (DMRs). When phased BAMs are available, haplotype-specific methylation metrics are incorporated to highlight allelic effects. A quantitative prioritization framework ranks events based on quality, magnitude, allelic imbalance, and regulatory context. Results are provided as annotated region files, cohort-wide z-score matrices, and interactive genome browser reports to facilitate interpretation.

## METAFORA Methods

METAFORA is a statistical framework for detecting and annotating methylation outlier regions from long-read CpG methylation data, integrating population reference modeling, hidden factor adjustment, and functional annotation, all while accounting for uncertainty from variable depth sequencing. **Figure 1** provides an overview of the computational workflow. We describe each step in detail along with input and output files below:

**Figure 1:**
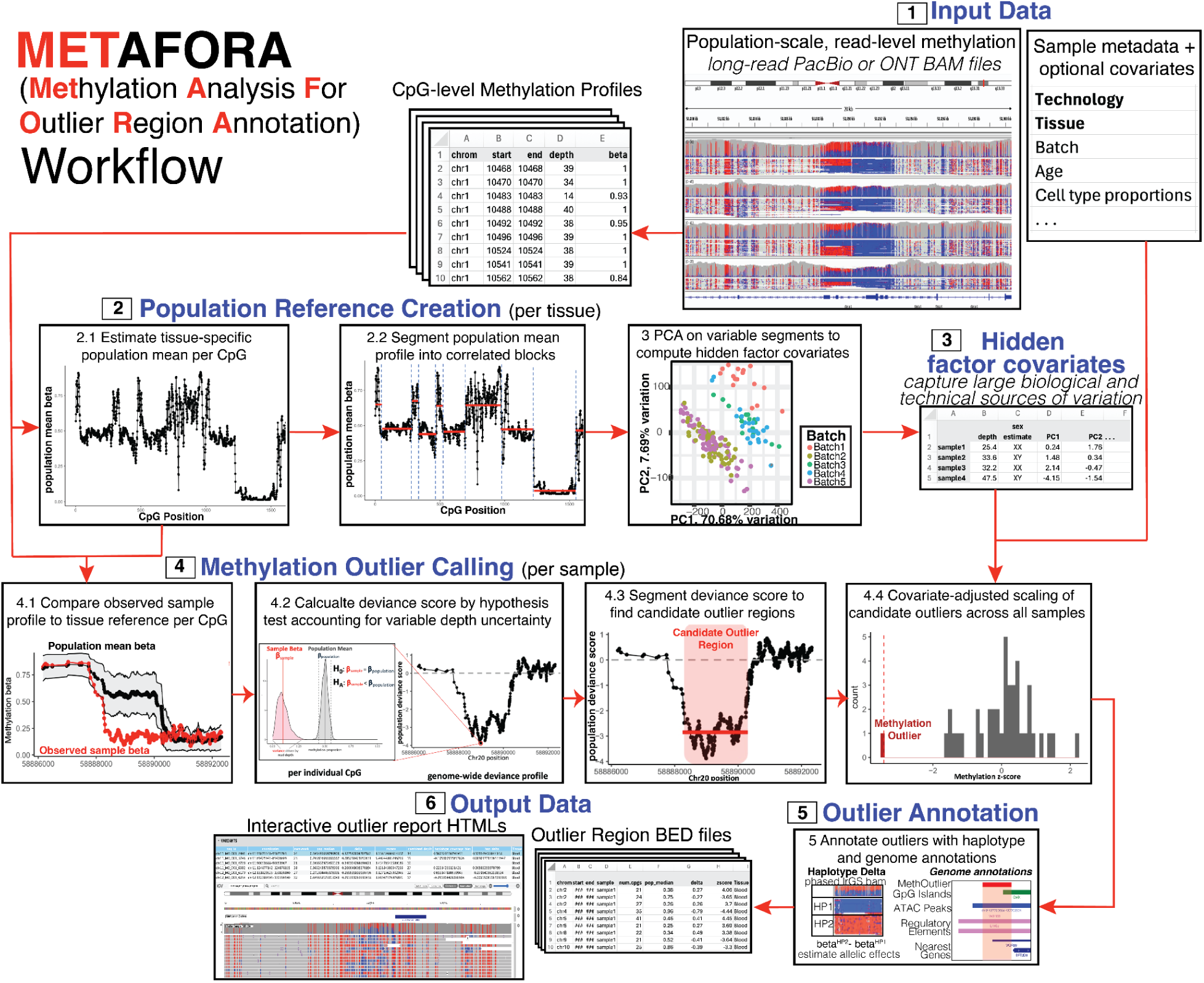
Metafora Workflow. In brief, lrGS bam and metadata is supplied as input data to METAFORA. CpG-level methylation profiles are aggregated to create tissue-specific reference of expected methylation and analyzed to find global hidden factors influencing genome-wide methylation. Individual sample-level methylation is compared to population reference to compute a deviance score at each CpG, which is then segmented into candidate outliers. Covariate-adjusted z-scores are then called over these candidate regions and thresholded to find methylation outliers. Outliers are annotated with genome tracks and haplotype information and output as a outlier region BED file and interactive IGV HTML report.

## 1 Input data and formatting

METAFORA expects aligned long-read BAM files containing methylation tags. For ONT data, this typically involves 5mC CpG methylation calls generated during Guppy or Dorado base calling; for PacBio, standard methylation calling from the jasmine pipeline is recommended (30). Users must supply sufficient samples from the same tissue type to reliably estimate expected methylation levels for outlier detection; we recommend a minimum of ∼30 samples per tissue to estimate robust z-scores. In addition to the BAM files, a sample metadata table is required, specifying the tissue of origin and sequencing platform (PacBio or ONT). An optional covariates file can also be provided. METAFORA will automatically estimate global hidden factor principal components (PCs) for correction during outlier calling (see Section 2.3), but users may also supply additional covariates to adjust for biological or technical factors, including batch variables, age, sex, or cell type proportions for instance.

From the long-read BAM files, METAFORA uses standard ONT and PacBio tools to aggregate methylation values across reads, generating CpG-level methylation profiles, modBam2Bed for ONT and pb-CpG-tools for Pacbio (31,32). For each CpG, the total read depth and methylation β-value (the proportion of methylated reads) are computed. These files are then sorted and indexed for efficient downstream retrieval. If 5hmC methylation is called during ONT basecalling, it is treated as an additional modified base, analogous to how it is handled in standard bisulfite sequencing. If an input BAM has been phased (as indicated in the metadata table), METAFORA will also generate haplotype-specific CpG methylation profiles, reporting β-values separately for HP1 and HP2.

Although METAFORA is designed for lrGS BAM files, it can also process precomputed methylation profiles if they are provided as properly formatted and indexed BED files containing read depth and β-values for each CpG. This enables compatibility with methylation calls from other tools (e.g., Nanopolish, f5c) or from alternative assay types such as bisulfite sequencing.

## 2 Creating population reference

To detect methylation outliers, METAFORA first builds a tissue-specific population reference that defines expected methylation levels at each CpG site.

### 2.1 Aggregating tissue methylation profiles

To construct this reference, METAFORA aggregates methylation profiles from all samples sequenced from the same tissue. Two matrices are generated: one containing β-values per CpG per sample, and another containing sequencing depth per CpG per sample. CpG sites with a median depth <5 across all samples are filtered out. The β-value and depth matrices are then combined to compute the mean proportion of methylated reads at each CpG. For memory efficiency, these calculations are performed separately for each chromosome chunk.

### 2.2 Segmentation of population reference

A key challenge of lrGS based methylation calling is that the breadth of individual CpG sites that are covered in a typical genome is greater than 36 million. Operating at single-CpG resolution across such extreme dimensionality results in prohibitive memory and time requirements and unstable estimation. To reduce dimensionality and improve estimation precision, the tissue-level population mean profile is segmented into correlated blocks using a change-point algorithm (fastseg) (33). Before segmentation, chromosomes are first chunked into distinct regions based on CpG density and each chunk is segmented independently to ensure that distant CpGs greater than 1kb apart will never be placed in the same segment. Also large segments composing more than 400 CpGs are broken up into smaller segments of at most 200 CpGs. Summarizing methylation over segments aggregates information across CpGs, mitigating the higher uncertainty caused by the lower coverage of lrGS compared with array-based assays. By averaging methylation and depth over correlated CpGs, segment-level methylation estimates achieve greater effective depth and reduced variance. Additionally, these segments capture biologically-relevant changes in methylation levels that are often observed in important regulatory sequences such as promoters and CpG islands or open chromatin regions.

## 3 Estimating hidden factor covariates

Global sources of biological and technical variation can obscure detection of cis-acting outliers (34); therefore, METAFORA estimates hidden factors from variable methylation segments and includes them as covariates to increase power for outlier detection.

### 3.1 Identifying variable methylation segments

To identify variable segments to input to the PCA step, segment β-values are transformed as: *Transformed = logit(1 - | beta - 0.5 |)*. This transformation forces symmetry about 0.5, mapping stable methylation states at either extreme (β≈0 or 1) to transformed values near zero, while more variable segments with mean β-values near 0.5 map to larger positive values. Then for each segment, the mean and variance of this transformed value are calculated across all samples, and the coefficient of variation squared (CV²) is computed. A LOESS curve is fit to the mean–CV² relationship, and residuals are calculated. The top 25% of segments by residual are defined as variable methylation segments, representing segments whose observed variability is greater than expected given that segment’s mean.

In addition to hidden factor estimation, these variable segment matrices can be used for downstream analyses such as differential methylation or QTL mapping. In another study, we applied this output to map methylation SV-QTLs in a large ONT cohort (35).

### 3.2 PCA of variable methylation segments

Principal component analysis (PCA) is performed on scaled M-value–transformed β-matrices from the variable segments. The optimal number of PCs is determined using the Gavish–Donoho random matrix theory threshold (36). These PCs can be used to explore batch effects and other sources of variation. METAFORA automatically generates PC plots colored by user-specified batch variables. Across datasets, sequencing depth often emerges as the largest driver of variation (see **Fig. 3F**); therefore, median depth is also included as a covariate during outlier calling. These PCs capture global sources of methylation variation–such as age, tissue cell type composition, and batch effects–and can improve power to detect local changes when properly corrected for (37).

### 3.3 Automated global outlier detection

METAFORA optionally detects and removes global outlier samples prior to outlier calling. Pairwise correlations between segment-level methylation profiles are calculated, and for each sample the mean correlation to all others is computed. This represents a similarity score for each sample to the rest of the cohort. In typical same-tissue datasets, mean pairwise correlations are centered ∼0.98; samples with substantially lower correlations are flagged as global outliers and removed. These often represent different tissue types, library preparations, or individuals with globally dysregulated methylation. The latter may be of interest in clinical contexts, and further investigation of global outliers is warranted. Recent work shows global outliers of splicing aided in rare disease diagnoses (38). Global outliers can also be identified from PCA plots as samples that do not cluster with the main cohort. Automated removal is optional and can be disabled in the workflow configuration to force outlier calling on all samples.

### 3.4 Automated sex chromosome inference

To account for sex-chromosome–associated methylation differences—particularly on the X-chromosome which is inactivated in XX individuals—METAFORA infers sex chromosome copy number from coverage data. Median coverage for chrX and chrY is normalized by genome-wide coverage and samples clustered into XX or XY groups. K-means clustering is performed with K=2, and cluster assignment probabilities assigned using a multivariate normal model of normalized X/Y coverage, with ambiguous cases (e.g., possible sex chromosome aneuploidy) labeled as such and excluded. Estimated sex is then included as an additional covariate, along with depth and hidden factor PCs, in the outlier calling step.

## 4 Outlier segmentation and calling

To identify methylation outliers, METAFORA compares each sample’s CpG profile against a tissue-matched population reference. Deviance scores quantify the probability that an observed sample deviates from the reference expectation given coverage uncertainty, and contiguous regions of deviance are segmented and filtered to yield candidate outliers.

### 4.1 Calculating deviance scores

At each CpG, METAFORA performs a beta distribution hypothesis test comparing the observed sample beta value to the expected population mean. Beta values are Laplace-corrected with pseudocounts to avoid boundary estimates at 0 or 1. A null distribution centered on the reference mean is parameterized by observed sequencing depth, with higher depth producing tighter variance. The cumulative probability of the observed sample beta is converted into a two-sided p-value, then transformed into a z-score for interpretability. Positive deviance scores represent hypermethylation, negative values hypomethylation, and values near zero signal agreement with the reference. To avoid inflated and infinite values from very deeply sequenced sites, coverage is capped at a user-supplied parameter (default 30 reads) and absolute deviance scores are winsorized at a depth-dependent threshold derived from the 10th percentile of simulated extreme outlier scores (delta = 1.0) (Supplemental figure **1A**).

### 4.2 Segmentation of deviance scores

The genome-wide deviance profile is segmented in parallel by chromosome. Profiles are first partitioned into blocks separated by >1 kb CpG gaps, then segmented within blocks using *fastseg* change-point detection. Two parameters guide segmentation: the minimum segment size and *median_seg_z*, a depth-adjusted threshold controlling sensitivity. *median_seg_z* is learned from simulations of outliers (delta = 0.25) across depths, using the 10th quantile deviance score as a cutoff to ensure ≥90% power at detecting outliers with deltas of at least 0.25. For each block, METAFORA computes the median depth and sets an appropriate *median_seg_z* via a fitted linear model (Supplemental figure **1B**). Mean deviance scores across candidate segments must exceed this threshold and meet the minimum CpG size.

### 4.3 Covariate-adjusted outlier calling

To call true outliers, METAFORA aggregates methylation across CpGs within each candidate segment for all samples, forming a sample × segment matrix. Beta values are transformed to M-values and residualized against user-supplied covariates and hidden factors inferred in step 2.3 using *edgeR*’s *correctBatchEffects* (*39*). This residualization corrects beta values for global sources of methylation changes, such as age, sex, and platform- or software-specific biases, increasing power to detect local changes. Residuals are then scaled and converted to z-scores. Segments where a sample’s absolute z-score exceeds a default threshold of 3 are defined as outliers. For each outlier, METAFORA reports the outlier delta (difference between sample and population median) and applies optional magnitude filters. Final outputs include a BED file of outlier coordinates with outlier statistics, plus the full sample × segment z-score matrix.

### 4.4 Joint outlier calling

Because METAFORA initially identifies outliers at the individual-sample level, the same underlying molecular event present in multiple samples may be represented by slightly different segment boundaries across individuals. To define a consistent cohort-wide set of events, individually detected outlier segments are first overlapped and merged across samples using *plyranges*. The resulting unified segment definitions are then used to reconstruct a cohort-level sample × segment matrix, and covariate adjustment, scaling, and z-score calculation are repeated as described in Section 4.3.

This procedure yields a harmonized joint outlier set in which segment boundaries are consistent across samples, facilitating direct comparison of outlier magnitude and recurrence across the cohort. METAFORA outputs both the individual-level z-score matrix and the jointly recalled matrix, enabling users to interrogate sample-specific deviations as well as shared events.

### 4.5 Sex chromosome outliers

Outliers on sex chromosomes are called using sex-stratified references to account for XX/XY differences in copy number and methylation. Cohorts are split by inferred sex, population means recalculated for chrX and chrY within each sex, and deviance scores recomputed within each group. The remaining steps mirror the autosomal pipeline, yet only include samples from the sex of the input sample.

All together, these steps define a robust pipeline that transforms CpG-level deviations into covariate-adjusted, segment-level outlier calls, while controlling for sequencing depth, noise, and sex-specific effects.

## 5 Annotation and prioritization of outlier regions

### 5.1 Outlier Annotation

After calling outlier regions per sample, METAFORA annotates each region using user-supplied genomic tracks to aid functional interpretation. By default, annotations include gene models, CpG islands, open chromatin (ATAC-seq peaks), and predicted Activity-by-Contact (ABC) enhancers with target genes (40), and curated coordinates of imprinting disorder differentially methylated regions (DMRs). For every outlier, METAFORA reports overlapping genes, whether the region intersects a CpG island, and any overlaps with regulatory elements (ATAC peaks or ABC enhancers) or imprinting DMRs. These annotations link non-coding methylation outliers to putative target genes and indicate whether outliers occur within established regulatory sequence. Outliers and nearby features can be explored interactively in an IGV HTML report generated for each sample (41).

Although phasing information is not used in calling outliers, METAFORA can add haplotype annotations when phased lrGS BAMs are provided. From phased reads, METAFORA computes haplotype-specific methylation profiles and intersects them with outlier coordinates. For each outlier it reports (i) haplotype-specific methylation proportions and coverage (HP1, HP2), (ii) the haplotype methylation difference (haplotype delta) and (iii) haplotype coverage bias (log ratio of HP1 to HP2 coverage). Large haplotype deltas highlights outliers with potential allelic effects—useful for interpreting cis-acting rare variants that would affect only a single haplotype—whereas outliers without allelic imbalance are more consistent with global influences (e.g. cell-type composition or technical noise). Haplotype delta can therefore be useful to filter outliers. Coverage-bias annotations also help flag false positives in imprinting contexts, where severe haplotype imbalance can artifactually mimic outliers.

### 5.2 Outlier Prioritization

To prioritize functionally or clinically relevant events, METAFORA implements a point-based scoring framework. Outliers receive points based on predefined criteria reflecting data quality, effect size, allelic specificity, and regulatory context. Quality metrics include minimum median sample depth (≥10 reads) and balanced haplotype coverage. Magnitude criteria include thresholds for absolute outlier delta, and allelic criteria include thresholds for absolute haplotype delta (Supplemental figure **2**).

Outliers also receive points for overlapping functionally relevant genomic annotations supplied by the user. By default, these include CpG islands, PBMC ATAC peaks, ABC enhancers, and imprinting disorder DMRs. Each overlap contributes a user-configurable weight. By default, imprinting disorder DMRs are assigned a higher weight (10 points) to emphasize well-established clinically relevant regions, whereas other annotations contribute one point.

Following scoring, outliers are filtered to those meeting a minimum score threshold (default ≥3 points) and ranked by prioritization score and absolute z-score. Highly ranked events are visualized in the METAFORA IGV HTML reports to enable efficient manual inspection and curation. This prioritization strategy focuses reporting on high-confidence, functionally relevant events, reducing report size and improving rendering efficiency compared to visualizing all detected outliers.

In summary, METAFORA helps functionally interpret outlier regions by systematically annotating nearby genes, CpG density, regulatory overlap, and (optionally) allelic effects and imbalance, and by providing an interactive prioritization and IGV report for rapid review.

## 6 Output data

METAFORA produces annotated outlier regions, a z-score matrix for downstream analysis, and interactive reports for rapid review.

### Primary outputs

● **Per-sample annotated BED** of outlier regions. For each outlier region METAFORA reports genomic coordinates, length, CpG count, coverage, population median beta, mean deviance score (from segmentation), sample z-score, outlier delta, and (when a phased bam is provided) additional haplotype metrics: HP1/HP2 methylation, HP1/HP2 coverage, haplotype beta difference, and coverage bias (log ratio HP1/HP2).
● **Outlier z-score matrix** (segments × samples) containing the covariate-adjusted methylation z-scores across the whole cohort for outliers identified in that sample and a cohort-level joint called matrix containing merged coordinates across all outliers identified.

### Interactive review

- ● **Per-sample IGV HTML report** visualizing a table of top prioritized outliers with overlaid annotations (genes, CpG islands, ATAC-seq peaks, ABC enhancers/targets) to facilitate manual curation and outlier visualizations.

### Quality control plots and summaries

● During reference construction, METAFORA outputs QC plots comparing median methylation across user-defined batches and cohort-wide correlation diagnostics.
● After outlier calling, METAFORA provides cohort outlier summaries: outlier counts by individual (stratified by batch, tissue, autosomes vs sex chromosomes) and distributions of segment length, CpG density, and outlier betas.

Overall, METAFORA is a depth-aware framework for detecting and annotating methylation outlier regions from lrGS CpG data: it builds a tissue-specific population reference, models uncertainty, and adjusts for hidden and user-supplied covariates. The pipeline computes CpG-level deviance scores, segments contiguous deviations, and calls covariate-adjusted outliers (including sex-stratified and optional haplotype-aware analyses). Outputs include per-sample annotated BEDs with summary statistics (and haplotype metrics when available), a cohort-wide z-score matrix, interactive IGV reports, and QC summaries.

## Results

### Benchmarking METAFORA on simulated data

We benchmarked METAFORA using simulated methylation outliers generated under a range of parameters (see Supplemental Methods). Outliers were created by sampling random reference sequences and introducing spike-ins defined by (i) population median beta, (ii) delta, outlier deviation from the median, (iii) cohort size (number of samples), (iv) number of CpGs in outlier segment, (v) average sequencing depth (vi) Gaussian noise added to beta values to mimic sequencing error and biological and technical variability. For each parameter combination, 100 outliers were simulated and METAFORA was evaluated based on sensitivity (≥50% reciprocal overlap with truth) and false positive rate.

### Sensitivity and power

METAFORA achieved high power across most conditions but struggled with small deltas (≤0.2) and low sequencing depth (**Fig. 2A**). At 5× depth, outliers were detectable only at large effect sizes (d ≥ 0.7). At 20× depth and above, sensitivity was near-perfect for d ≥ 0.3, while only 30× depth consistently reached >50% power at d = 0.2. Increased noise further reduced power, particularly for short outliers (5–10 CpGs), while larger segments were able to effectively average out noise effects. Even without noise, very short segments remained more difficult to recover, likely reflecting limitations of change-point segmentation for small intervals (**Fig 2B**).

**Figure 2:**
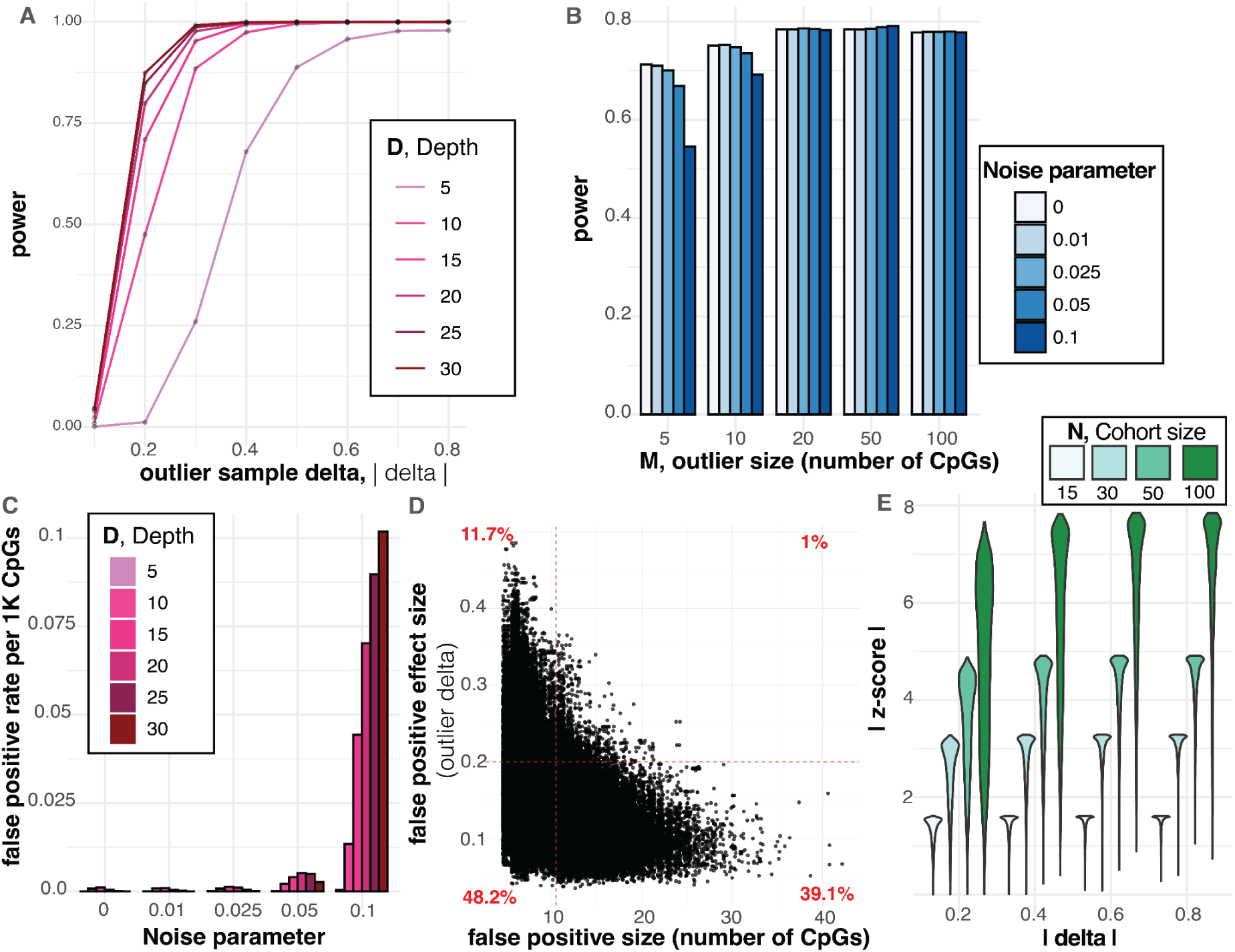
METAFORA benchmark on simulated methylation outliers. **A** Sensitivity of METAFORA detecting simulated outliers based on varying sequencing depth and outlier delta (difference of outlier methylation proportion compared to median population). **B** Sensitivity of METAFORA detecting simulated outliers based on outlier size (number of contiguous CpGS in outlier segment) and noise parameter (variance of simulated noise added to beta estimates to mimic sequencing/methylation calling error). **C** Number of false positive outlier segments called in background methylation regions as a function of increasing noise parameter and sequencing coverage. **D** Characteristics of false positive outlier regions in terms of methylation size (number of CpGs) and outlier delta. **E Z-score** of simulated outlier segments as a function of outlier delta and cohort size, showing increasing cohort size increases estimated z-scores for true outliers.

### False positives

False discovery rates were primarily influenced by noise, e, and, to a lesser extent, sequencing depth (**Fig. 2C**). Higher depths reduced sampling variance of beta estimates allowing noise to have an inflated effect when calculating deviance scores. CpGs that vary from median due to noise can then be erroneously segmented as deviant. While false positives were prevalent at high noise and depth levels, we found these erroneous calls tended to be small (< 10 CpGs) or had minimal delta values (< 0.2). Applying these filters removed 99.95% of false positives, while having a small impact on true outliers (**Fig. 2D**).

### Cohort Size

Increasing the number of samples in our simulations strongly increased z-scores of detected outliers (**Fig. 2E**). Higher z-score enables better stratification of true outliers from noise, and underscores the importance of having a large cohort as input to METAFORA. The increased confidence from larger cohort sizes was particularly helpful at rescuing weak outliers (d = 0.1**–**0.2) that would have otherwise fallen below significance threshold. For d = 0.1, for instance, a cohort of 100 detected nearly twice as many true outliers as a cohort of 15. Similarly, for d = 0.2, a cohort of 100 improved detection of outliers to 84.4% compared to 81% with a cohort of 15.

Together, these simulations demonstrate METAFORA is robust and well-powered to detect outliers across a wide range of parameters yet highlight high noise or low-depth sequencing as main limitations. We optimized METAFORA parameters and post-filtering strategies according to these results to minimize false positive calls.

### Comparison of sequencing platform methylation profiles

Having established METAFORA’s sensitivity in simulations, we next evaluated it on real lrGS data from the Undiagnosed Diseases Network clinical site at Stanford and the Genomics Research to Elucidate the Genetics of Rare disease (GREGoR) R02 release. This cohort of long-read genomes spans multiple technologies and chemistries—Oxford Nanopore Technologies (ONT) R9.4 and R10.4, and PacBio Revio—and multiple sequencing sites (Broad, GSS/Stanford, BCM, PMGRC, UW), enabling assessment of platform differences and potential site effects (42).

Our dataset comprised 551 genomes (77 R9, 98 R10, 376 PacBio) from blood (n=503) and fibroblast (n=48). Sequencing metrics varied widely across platforms and sites. PacBio achieved high coverage (>35× median) with more constrained read-length N50 (median 17.2 kb). ONT R9 (primarily from GSS/UDN) had lower coverage (median ∼25×) with a broad read-length range (median ∼20 kb). ONT R10 matched PacBio coverage (median ∼38×) but with longer reads (median 33.7 kb; some N50 up to 50 kb and above) (**Fig. 3A**). Given our simulation results showing that increasing sample size improves detection power and yields higher absolute outlier z-scores, our goal was to combine samples across platforms to analyze as large a cohort as possible. However, because platform-specific artifacts could bias analyses, we first compared methylation profiles across technologies and sites to determine whether joint modeling is appropriate.

**Figure 3:**
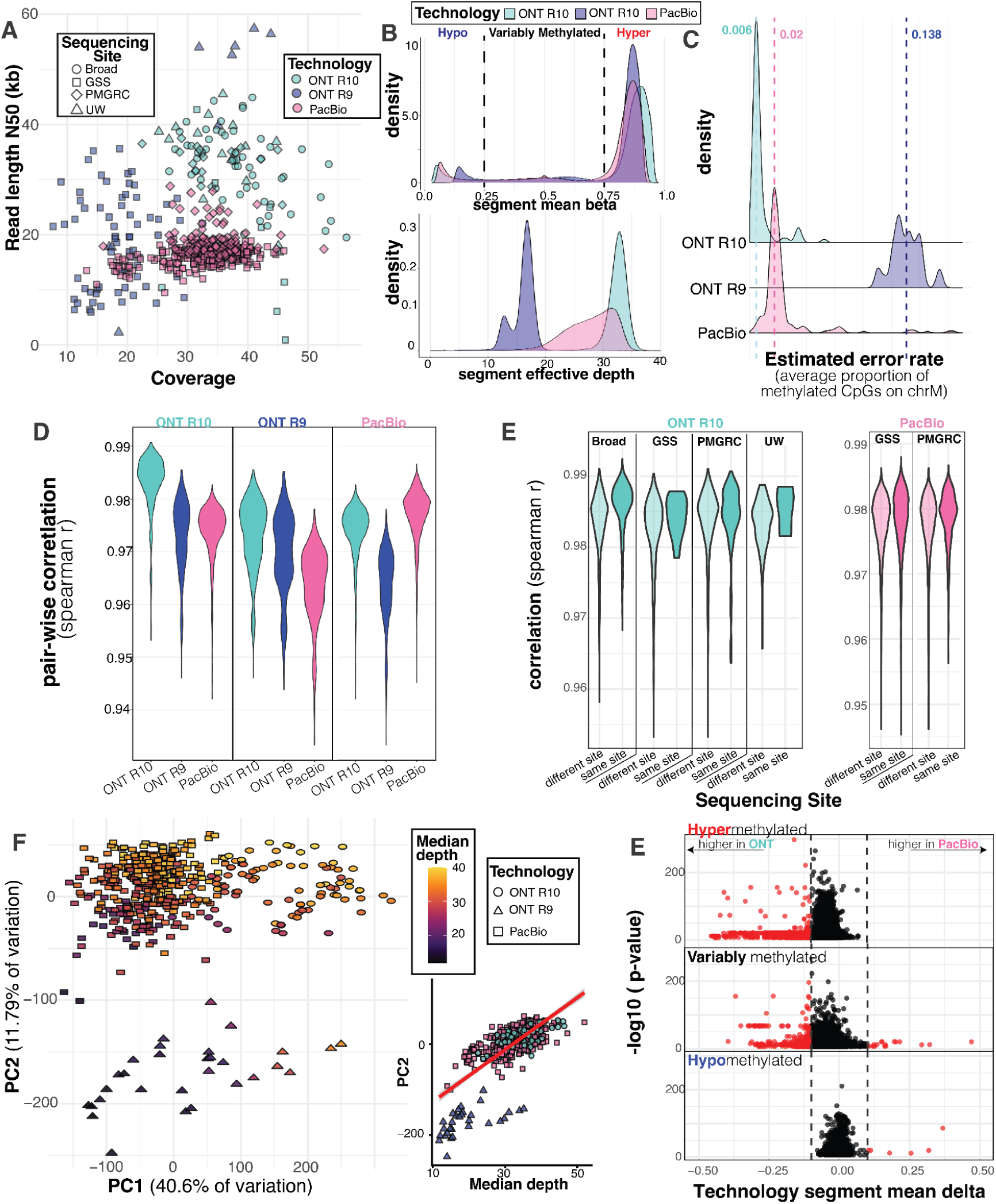
Technology comparison of genome-wide CpG methylation detection. **A** Scatter plot displaying sequencing metrics of read length and coverage across different technologies and sites from UDN/GREGoR lrGS data. **B** Distribution of the median methylation profile and effective depth of segmented genome bins across three different platforms. **C** Distribution of estimated error rate for each technology by averaging the number of erroneous methylated calls on chrM which is effectively unmethylated. **D** Distribution of pair-wise sample correlations stratified by whether samples were sequenced from the same or different platforms. **E** Distribution of pair-wise sample correlations within each platform, stratified by whether sample was sequenced at same or different site. **F** First two principal components of the methylation segment matrix show clustering by sequencing platform and stratification with respect to depth. **G** P-value and observed sample delta from linear modeling of methylation by sequencing platform for all genome segments. Thresholds at absolute delta of 0.1 and red points exceeding them represent substantively large changes in methylation profile.

### Comparison of segment methylation profiles

To achieve this, we constructed a joint population reference using all samples, segmented the mean profile, and summarized per-sample methylation within segments; platform-specific median profiles were then computed. Segment mean beta distributions were consistently bi-modal (reflecting hypo- and hyper-methylated states) as expected and followed similar distributions across platforms albeit with some variation (**Fig. 3B**). The ONT R10 hypermethylated peak was closer to one with tighter variance than PacBio or R9, and its hypomethylated peak was also closer to zero; PacBio and especially R9 showed higher beta in the hypo peak and lower beta in the hyper peak.

To explore what could be mediating shifting beta distributions, we also compared the **effective depth** of each segment: the average number of confidently called reads per segment. This metric captures not only total depth but also mappability and the percent of reads that could be confidently called due to per-read methylation accuracy. ONT R10 had the highest effective depth (centered ∼33×), while PacBio had a significantly left-skewed tail, and R9 was lowest (**Fig. 3B**). Lower effective depth increases the influence of pseudocount correction and sampling variance, pushing β estimates away from 0/1; likely accounting for the broader tails observed for PacBio and R9.

Defining a quantitative measure of noise or error rate in methylation calling across platforms is difficult because there is no genomic truth set of known methylation in the nuclear genome. We, therefore, estimated a per-sample proxy for methylation error using the mitochondrial genome, which is effectively unmethylated at CpG sites as a result of its prokaryotic origin (43,44). After restricting to primary alignments (to avoid NUMT artifacts), we calculated the fraction of methylated CpGs on chrM to yield platform-specific error proxies: ONT R10 median **0.6%**, PacBio **∼2%**, ONT R9 **13.8%** (**Fig. 3C**). These estimates are largely concordant with the effective-depth differences and help explain the attenuated extremes for PacBio and R9.

### Pairwise correlation across platforms and sequencing sites

In addition to comparing the median profile from each technology, we also computed pairwise correlations across segmented methylation profiles in whole blood samples, removing global outliers with mean pairwise r < 0.95 (mostly low-depth R9 and several PBMC samples). Overall concordance was high (median Spearman r = **0.976**) even across disparate sequencing platforms. Same-technology pairs correlated slightly more than cross-technology pairs: ONT R10–R10 mean r = **0.984** vs R10–R9 **0.975** and R10–PacBio **0.974**; with similar trends observed for other platforms (**Fig. 3D**). Interestingly, we observed ONT R10 had the highest intraplatform correlation compared to PacBio (mean r = 0.978) and ONT R9 (mean r = 0.971), mirroring observed effective depth and estimated error rates.

To investigate potential sequencing site-specific effects, we compared within-technology correlations across different sequencing centers. Differences between same-site and cross-site pairs were minimal (R10 mean-correlation difference ≈ 0.0024; PacBio ≈ 0.0004), indicating little batch structure attributable to site (**Fig. 3E**). This contrasts with other functional omics (e.g., RNA-seq or proteomics), where batch effects can be more prevalent (45); in our data, genome-wide CpG methylation appears comparatively resilient to batch effects, consistent with its role as a stable epigenetic mark.

### Sequencing depth is major source of variation in methylation profiles

We performed principal component analysis on the methylation segment matrix to estimate global sources of variation. Visualizing the first two PCs displayed clear clustering based on sequencing technology. PC1 captured mostly platform-specific differences stratifying R10 and PacBio samples, while PC2 stratified samples within each platform by sequencing depth.

Interestingly, both principal components significantly stratified samples according to depth, particularly for PC2 (depth-PC1 correlation: 0.36, depth-PC2 correlation: 0.74) (**Fig. 3F**). This reinforces the finding that sequencing depth is a major determinant of the variability of methylation segments and mitigates a lot of the technology-specific biases observed between the different platforms.

To assess the specific segments that display platform-based differences, we called differential methylation between R10 ONT and PacBio samples. We transformed and scaled beta values into M-values and ran a linear regression predicting segment methylation by sequencing platform, adding depth as a covariate to control for any effect from sequencing coverage. Our differential methylation model revealed that most segments did have significant differences between the two platforms, but observed effect sizes tended to be small in magnitude.

Thresholding differentially methylated segments based on observed median beta difference of at least 0.1 between ONT and PacBio, revealed only 2.3% of segments as substantially technology-biased (**Fig. 3G**). Most biased segments lay in hypermethylated regions (median beta > 0.75) where methylation was lower in PacBio compared to ONT, consistent with a lower effective depth and higher error; fewer differences were observed in variably methylated segments, and only rare differences in hypomethylated regions.

All together, our comparisons establish that platform-specific biases in genome-wide CpG methylation are detectable but generally modest. We find methylation profiles to be highly concordant across technologies and sites. Effective depth and error rate largely explain the small observed differences where they occur. These results support joint analyses across platforms provided that technology and depth are modeled as covariates (and/or addressed via hidden-factor adjustment).

### METAFORA detects multi-platform methylation outliers

We ran METAFORA on the combined GREGoR/UDN cohort to identify multi-platform methylation outliers using conservative thresholds chosen to limit noise-driven false positives (minimum segment size = 20 CpGs, minimum absolute delta > 0.3, and minimum absolute z-score > 3).

### Outlier burden per genome

Across the cohort, METAFORA identified 32,808 outlier regions (26,816 in blood; 5,992 in fibroblast), with a loss of methylation being slightly more common (58.8% hypomethylated vs 41.2**%** hypermethylated). Outliers detected in blood also exhibited higher absolute z-scores than those in fibroblasts, consistent with the larger blood cohort improving power (**Fig. 4A**). Per sample, the median autosomal burden of large outliers (>30 CpGs) was 17. Counts were broadly concordant across platforms, though PacBio genomes showed a higher per-genome burden than ONT (PacBio median = 21, ONT R9 median = 13, ONT R10 median = 6). Beyond autosomes, METAFORA called 1,221 sex-chromosome outliers using the sex-stratified pipeline. We observed a median of 3 outliers per genome on chrX and, in XY individuals, a median of 1 outlier on chrY (**Fig. 4B**). A majority of sex-chromosome outliers arose from PacBio genomes.

**Figure 4.**
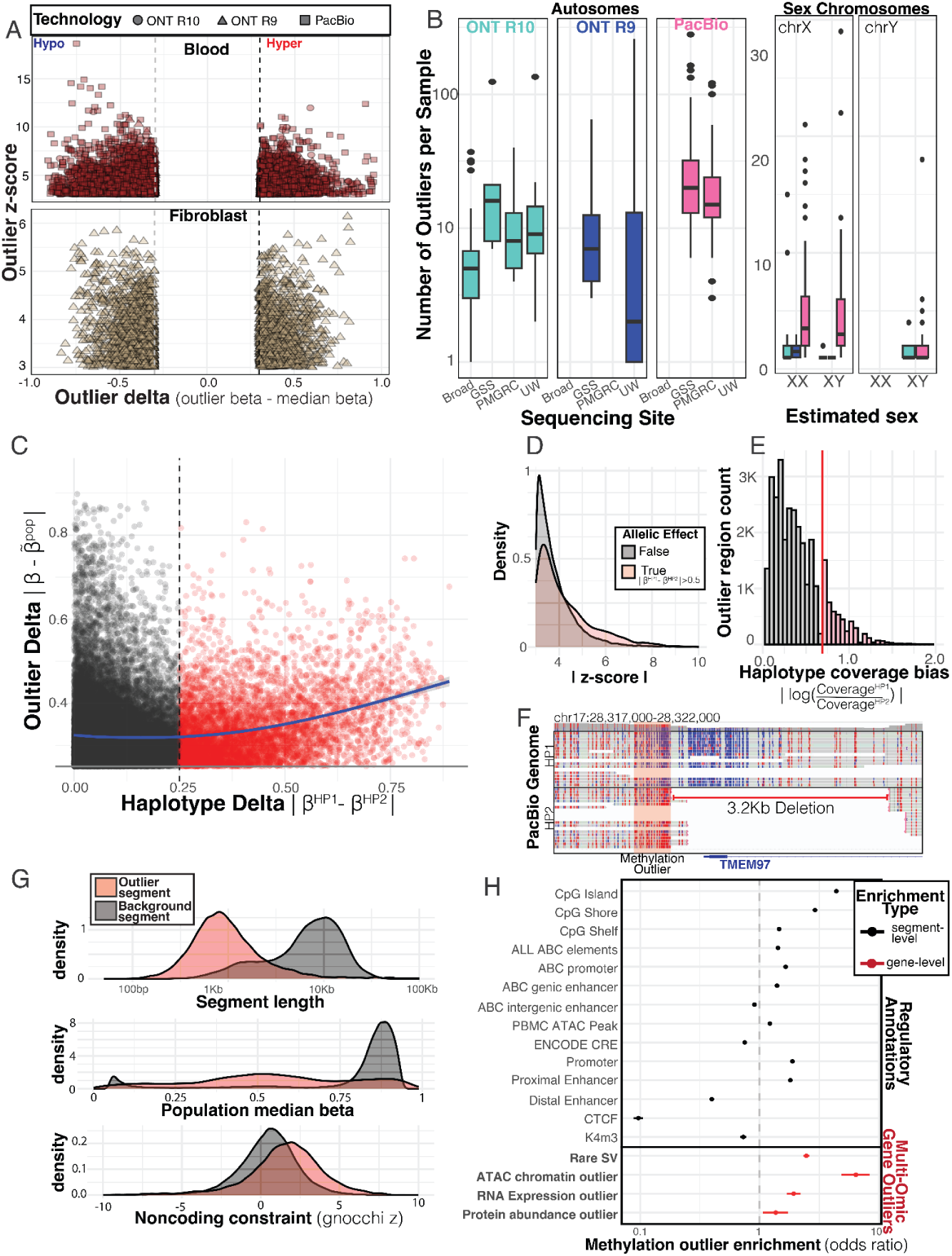
METAFORA identifies functionally important methylation outlier regions from multi-platform lrGS data. **A** Volcano plot of outlier effect size (beta delta) and outlier p-value across both blood and fibroblast. **B** count of outliers per genome across different sequencing platforms and sites on both the autosomes and sex chromosomes. **C** Scatter plot of methylation outlier regions from PacBio genomes by haplotype-specific allelic effects on x-axis with population delta on the y-axis, segments with large allelic-effects (haplotype delta > 0.25) are highlighted in red. **D** Z-score distribution showing shift in z-scores of large allelic-effect outliers (absolute haplotype delta > 0.5). **E** Histogram of haplotype coverage bias across PacBio methylation outliers, highlighting outliers where one haplotype had more than two-fold coverage compared to the other. **F** Methylation IGV read pile-up from a phased PacBio genome with heterozygous deletion of *TMEM97* promoter along with a proximal gain-of-methylation outlier on the same haplotype. **G** Distribution of outlier region length, median population beta, and noncoding constraint score for cohort outlier calls compared to background methylation segments from population mean profile segmentation. **H** Regulatory genome annotation enrichment of various tracks overlapping methylation outlier regions compared to background segments. Bottom plot displays gene-level enrichment of observing a gene within 10kb of methylation outlier also nearby a rare structural variant or multi-omic outlier in ATAC-seq, RNA-seq, or proteomics. Enrichment odds ratio and confidence interval from Fisher’s exact test is plotted.

### Phased long-read genomes reveal haplotype-specific outliers

We used PacBio genomes to explore whether methylation outliers often show haplotype-specific effects because this platform was consistently phased with the same workflow across different sequencing sites. For each outlier, METAFORA reported the methylation level and coverage for each haplotype, as well as the difference in methylation between them (the haplotype delta) and any coverage imbalance.

We found that 16.4% of outliers showed clear allelic effects, e.g. regions where one haplotype was substantially more methylated than the other (**Fig. 4C**). Outliers with strong haplotype differences also tended to deviate more strongly from the population median and had higher z-scores overall (**Fig. 4D**). This pattern supports the idea that these are true biological outliers rather than false positives, since noise would be expected to affect both haplotypes equally.

Filtering outliers to those with large haplotype delta reduces the number of outliers from 21 to only 4 per genome, easing the burden of manual curation. We also investigated coverage balance across haplotypes, where we found that many genomes showed skewed representation of one haplotype over the other. Coverage bias is usually a technical artifact from the imbalanced sampling of haplotypes during lower-depth sequencing, but could also signal real copy number changes. We found 16.2% of outliers had more than a two-fold difference in coverage between haplotypes (**Fig. 4E**). Such imbalance can complicate interpretation and detection of outliers, especially in regions subject to parent-of-origin effects, where skewed coverage mimics the effect of an outlier.

Manual inspection of outliers with large haplotype effects revealed that many haplotype-specific outliers coincide with nearby rare genetic variants, including upstream deletions, mobile element insertions, CGG repeat expansions, and CpG-rich VNTR insertions (Supplemental figure 3).

Systematically, we found that rare structural variants called from the lrGS data were 2.47 fold enriched near methylation outliers genes (p-value < 2.2e-16) (**Fig. 4H**). Case in point, we identified a gain-of-methylation outlier near the *TMEM97* promoter in a sample carrying a 3.2kb deletion spanning the promoter/5′ UTR. Phasing confirmed that the methylation outlier lies upstream of the deletion on the same haplotype, consistent with a cis-acting structural variant. In this scenario, the deletion likely removes part of the promoter and disrupts transcription initiation which in turn could alter local chromatin context and methylation state on the affected haplotype (**Fig. 4F**).

### Outlier characterization reveals their functional importance

We further sought to explore common characteristics of outliers and where they appear in the genome. We found that outlier regions are typically compact, spanning ∼100bp-1kb in length with a median of 28 CpGs. Compared to background methylation segments, we found outliers were also strongly enriched in variably methylated regions of the genome (16.5 times more likely to have population median beta between 0.25 and 0.75), which in whole-genome segmentation were relatively rare, yet make up the majority of our outlier regions (**Fig. 4G**). We tested if methylation outlier regions from blood also occurred in functionally important regulatory sequence, and found that outliers were enriched in CpG islands (odds ratio (OR) = 4.27), open chromatin ATAC peaks from PBMCs (OR=1.22), annotated promoters (OR=1.89), and Activity-by-contact (ABC) predicted regulatory elements (OR=1.43). Interestingly, outlier regions tended to be more enriched in proximal or intragenic regulatory regions compared to more distal enhancers (**Fig. 4H**). Further, compared to background methylation segments, outlier segments displayed higher noncoding variant constraint scores from Gnocchi (t value=57.1), again underscoring their functional importance (46).

We then assessed downstream impact using multi-omics data available in the same UDN and GREGoR samples, including ATAC-seq, RNA-seq, and proteomics. Genes within 10Kb of methylation outliers were significantly more likely to also have outlier signals in other omics layers, most strikingly in chromatin accessibility (OR=6.41, p-value=1.03e-28), but also in gene expression (OR = 1.93, p-value=3.7e-19) and protein abundance (OR = 1.37, p-value=0.011) (**Fig. 4H**). This cross-omic enrichment highlights that methylation outliers detected by METAFORA are not random events but instead mark regions with large regulatory effects that propagate through the molecular cascade, and in particular could tag aberrant changes in chromatin accessibility.

One example of a METAFORA methylation outlier region with strong multi-omic signal occurred in the promoter / 5’ UTR of the FAM193B gene. We identified an unaffected individual in GREGoR sequenced on PacBio who had a large 2,025bp insertion at this STR locus, corresponding to 675 additional CGG repeat copies. METAFORA identified a strong hypermethylation outlier in phase with this expanded haplotype (z-score = 6.68, |haplotype delta| = 0.785) (Fig. 5A,B). In matched multi-omic data from the same individual, the corresponding promoter-associated ATAC-seq peak showed markedly decreased chromatin accessibility (EpiOut *p-value*= 9.85e-7; log2 fold-change = −0.78), accompanied by strong underexpression of *FAM193B* in blood RNA-seq data (expression z-score = −4.04) (Fig. 5C,D). Together, these observations support coordinated epigenetic and transcriptional silencing of the expanded haplotype.

**Figure 5.**
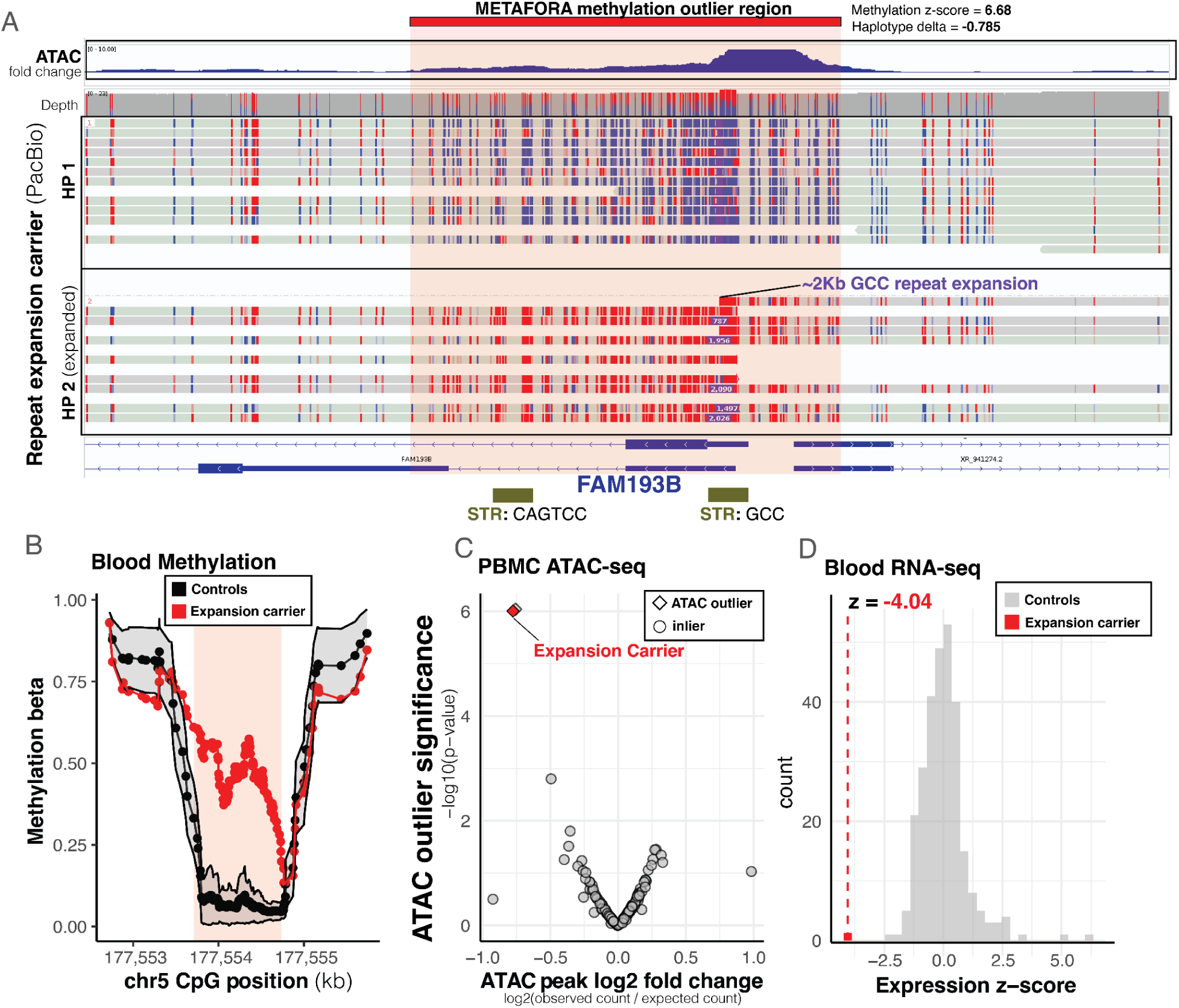
METAFORA tags multi-omic outlier at *FAM193B* across multiple layers of regulation. **A** IGV screenshot of PacBio genome where a rare 2.0Kb CGG repeat expansion was identified overlapping strong METAFORA hypermethylation outlier. ATAC-seq fold change signal from MACS2 also displayed from the same individual. Gene model as well as STRs from adotto tandem repeat catalog displayed below. **B** Methylation beta line plots show mean and standard deviation of population methylation proportions over the expanded outlier region compared to the observed methylation proportions from the *FAM193B* expansion sample. Red box marks outlier boundaries defined from segmentation of the deviance score. **C** ATAC-seq outlier fold change and p-value from EpiOut for the FAM193B promoter peak (chr5:177552709-177555736). Fold change on X-axis plots log2 ratio of the observed ATAC read counts to the expected read counts, and p-value on Y-axis measures significance of the observed accessibility outlier status. The sample colored in red represents the expansion carrier. Diamond points represent samples called as outliers by EpiOut. **D** Histogram of normalized expression z-scores of *FAM193B* from Blood RNA-seq data showing expansion carrier (in red) is a strong underexpression outlier.

Applying METAFORA on real-world data revealed that methylation outliers arise across platforms and tissues, frequently showing haplotype specificity and enrichment near rare variants. These regions are concentrated in regulatory and conserved sequence and often coincide with chromatin, expression, and protein outliers, highlighting their functional relevance and potential as markers of cis-regulatory disruption.associated with the expanded allele.

### Case study: METAFORA outliers aid in rare disease diagnosis

Since the UDN and GREGoR long-read genomes were derived from rare disease patients and family members, we asked whether methylation outliers could provide complementary information to support molecular diagnoses. Imprinting disorders provide a natural test case, as they are well-established syndromes caused by disrupted methylation at imprinted loci and should manifest as outliers in our framework.

We curated a list of 12 differentially methylated regions (DMRs) associated with known imprinting disorders (Table 1) (9) and intersected these with METAFORA outliers. This analysis identified 43 candidate outliers across 40 individuals. To prioritize the most clinically relevant outliers, we applied additional filtering steps, including requiring high read coverage to reduce false positives, manual inspection of read pileups to exclude cases driven by haplotype-specific coverage imbalance, and requiring phenotypic overlap between the proband and the expected imprinting disorder based on the direction of the methylation outlier (Fig. 6A). The vast majority of imprinting disorder outliers displayed haplotype imbalance, and filtering these drastically reduced the number of unaffected individuals represented among prioritized outliers. After applying these filters, three compelling candidate cases remained, involving methylation outliers at the imprinting regions **GNAS** and **KCNQ1OT1**, which were prioritized for further investigation.

**Figure 6:**
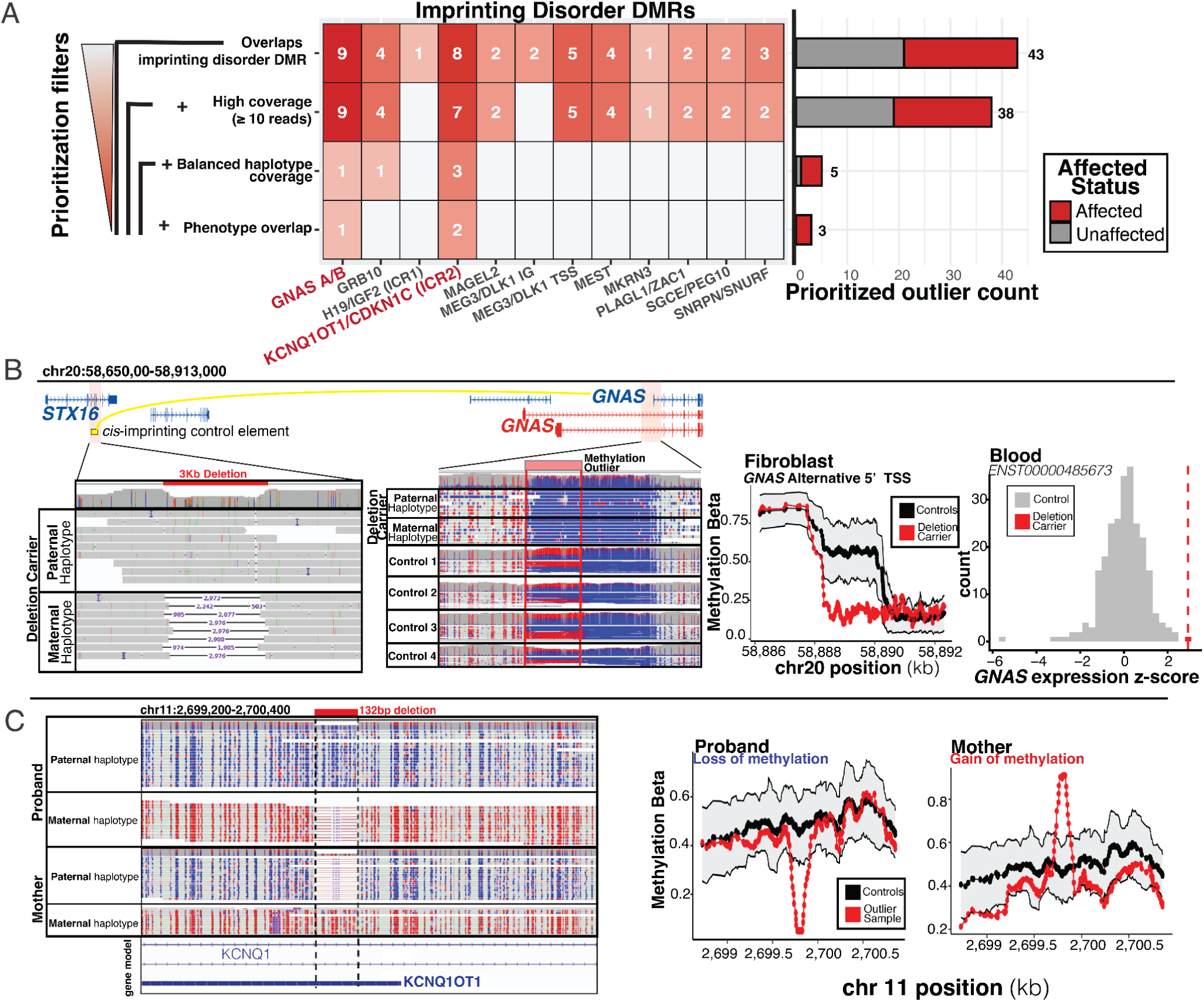
METAFORA outliers aid in diagnosis of imprinting disorders. **A** count of patients with prioritized imprinting disorder outlier, colored by affected status. Y-axis displays filtration schema that are progressively applied to filter down outliers, and x-axis of heatmap displays in which imprinting disorder regions these outliers were found. **B** Read pileup from ONT R9 data of a 3kb *STX16* deletion on maternal haplotype as well as loss-of-methylation outlier on same maternal haplotype in the *GNAS* gene for a UDN case. Control samples included in methylation pile-up to show expected imprinting methylation pattern of 50% methylation at this region. Methylation line plot shows the deletion carrier has significant loss-of-methylation compared to population mean and standard deviation. Histogram of *GNAS* expression from RNA-seq shows the patient is also an over-expression outlier with the highest z-score of the full cohort. **C** IGV read pile-up of PacBio data from a GREGoR family displaying a rare 132bp deletion in the *KCNQ1OT1* imprinting control loci leading to a loss-of-methylation outlier in proband and gain-of-methylation outlier in mother due to deletion occurring on differing parental chromosomes. Methylation beta line plots show mean and standard deviation of population methylation over the expanded outlier region compared to observed sample-level methylation.

**Table 1:** Imprinting disorder regions. Curated list of imprinting disorder DMRs were selected from a recent review (9) and visualized in IGV to determine which loci displayed reliable imprinting in blood that is detectable by lrGS. DMR coordinates were defined based on observed imprinting boundaries displaying allele-specific methylation across multiple samples.

| DMR coordinates | DMR Name | Expressed Allele | Outlier Direction | Associated Disease |
| --- | --- | --- | --- | --- |
| chr6:144,006,966-144,008,688 | PLAGL1/ZAC1 | Paternal Expressed | Loss of methylation | Transient Neonatal Diabetes Mellitus |
| chr7:50,781,054-50,783,474 | GRB10 | Maternal Expressed, but can be tissue/isoform dependent | Loss of methylation | Silver-Russell Syndrome |
| chr7:94,656,289-94,658,660 | SGCE/PEG10 | Paternal Expressed | Gain of methylation | Silver-Russell Syndrome |
| chr7:130,490,205-130,493,321 | MEST | Paternal Expressed | Gain of methylation | Silver-Russell Syndrome |
| chr11:1,997,714-2,003,214 | H19/IGF2 (ICR1) | Maternal Expressed (H19), Paternal Expressed (IGF2) | Gain of methylation | Beckwith-Wiedemann Syndrome |
|  |  |  | Loss of methylation | Silver-Russell Syndrome |
| chr11:2,698,696-2,700,861 | KCNQ1OT1/CDKN1C (ICR2) | Paternal Expressed (KCNQ1OT1), Maternal Expressed (CDKN1C) | Gain of methylation | Silver-Russell Syndrome |
|  |  |  | Loss of methylation | Beckwith-Wiedemann Syndrome |
| chr15:23,561,746-23,562,545 | MKRN3 | Paternal Expressed | Loss of methylation | Central Precocious Puberty |
| chr15:23,646,980-23,648,066 | MAGEL2 | Paternal Expressed | Gain of methylation | Schaaf-Yang Syndrome |
| chr15:24,954,757-24,957,340 | SNRPN/SNURF | Paternal Expressed | Gain of methylation | Prader-Willi Syndrome |
|  |  |  | Loss of methylation | Angelman Syndrome |
| chr14:100,809,270-100,811,430 | MEG3 IG-DMR (MEG3/DLK1) | Maternal Expressed (MEG3), Paternal Expressed (DLK1) | Gain of methylation | Kagami-Ogata Syndrome |
|  |  |  | Loss of methylation | Temple Syndrome |
| chr14:100,824,177-100,827,570 | MEG3-Promoter DMR | Maternal Expressed | Gain of methylation | Kagami-Ogata Syndrome |
|  |  |  | Loss of methylation | Temple Syndrome |
| chr20:58,888,073-58,890,209 | GNAS A/B DMR | Paternal Expressed | Gain of methylation | Pseudohypoparathyroidism Type 1A |
|  |  |  | Loss of methylation | Pseudohypoparathyroidism Type 1B |

### *STX16* deletion leads to loss of imprinting of *GNAS* gene

In the UDN, a proband with overgrowth and marked infantile-onset obesity (weight z-score at 17 months old was Z=6.98) was evaluated alongside a similarly affected sibling. Long-read ONT R9 sequencing of cultured fibroblasts from the proband identified a 3 kb deletion involving exons 4-6 of *STX16* (Chr20(GRCh38):g.58668511_58671488del) previously identified by short read genome sequencing as well as a loss-of-methylation outlier at an alternative *GNAS* promoter (delta = −0.49, z-score = −3.84) (**Fig. 6B**). The deletion has been reported in the literature in multiple individuals with pseudohypoparathyroidism type 1B (PHP1B) and is classified as pathogenic in ClinVar (Variation ID: 978043) (47–50). Normally, *GNAS* is silenced on the maternal allele and expressed only paternally, but deletion of *STX16* on the maternal haplotype is purported to disrupt a cis-imprinting control element, causing methylation loss at *GNAS* and aberrant biparental expression (51–53).

Long-read phasing confirmed that the *STX16* deletion and the methylation outlier co-occurred on the same maternal haplotype. Further, RNA-seq data from blood provided evidence the proband was an overexpression outlier for GNAS transcript (gene-level z-score=2.05, isoform-level z-score=2.93 for alternative 5’ start transcript *ENST00000485673*) consistent with aberrant biparental expression. Short-read sequencing of the family confirmed the deletion was also present in the affected sibling and inherited from the mother, who remained unaffected presumably because her deletion is present on the paternal chromosome. Together, these findings provide strong support for a diagnosis of PHP1B in the proband and sibling.

### KCNQ1OT1 microdeletion in overgrowth syndrome

In one GREGoR family, the proband presented with tall stature, developmental delay, and toe walking. Prior panel testing for overgrowth syndromes and Prader–Willi methylation testing were negative, and the patient was recruited to GREGoR for multi-omics evaluation. PacBio sequencing from whole blood and METAFORA identified a 120 bp methylation outlier (delta = −0.45, z-score = −7.5) in the *KCNQ1OT1* IC2 imprinting region, a locus implicated in Beckwith–Wiedemann syndrome (BWS) and Silver–Russell syndrome (SRS). The outlier was tagging a 132bp deletion present on the proband’s silenced maternal chromosome, leading to relative loss of methylation due to the lack of DNA, while imprinting in the rest of the locus remained intact. The same deletion was inherited from the mother, who carried it on her unmethylated paternal chromosome and was also called as a methylation outlier in the opposite direction, since the unmethylated copy was deleted (delta = 0.44, z-score = 5.8) (**Fig. 6C**).

This rare deletion has been reported to cause SRS growth-retardation symptoms when paternally inherited, through loss of function of the lncRNA KCNQ1OT1 and subsequent dysregulation of its target CDKN1C (54,55). Another case report, however, described maternal transmission of the deletion associated with BWS-like features, including overgrowth, cancer predisposition, and shortened Achilles tendon, consistent with the proband’s toe-walking and overgrowth phenotype and cautions interpretation of this variant as a variant of uncertain significance(56). The deletion also overlaps a structural variant reported in gnomAD (Chr11(GRCh38):g.2699748_2699865del; allele frequency of 0.05% in gnomAD v4.1.0), including several homozygous individuals. Although the proband’s phenotype partially overlaps reported presentations, the recurrence of this variant in population datasets suggests it is unlikely to be a fully penetrant cause of rare imprinting disorders. Nevertheless, this example illustrates how methylation outliers can highlight rare regulatory variants and prioritize candidate loci for further investigation.

Together, these case studies illustrate how METAFORA can surface clinically relevant methylation outliers in rare disease cohorts. By combining long-read sequencing, phasing, and multi-omics integration, the framework not only pinpointed known imprinting loci but also connected outliers to underlying structural variants, allelic effects, and downstream transcriptomic changes. Together, these findings demonstrate the potential of methylation outlier analysis to complement standard genetic testing and provide new avenues for diagnosis in unsolved rare disease cases.

## Discussion

Rare variant interpretation remains a central barrier in rare disease diagnosis, but incorporating genome-wide methylation signals is providing new opportunities, particularly for imprinting disorders and other epivariants. Here we introduce **METAFORA**, a software framework for detecting methylation outliers genome-wide from population long-read genome sequencing.

METAFORA models uncertainty from variable sequencing depth, adjusts for global hidden factors influencing methylation, and segments deviant methylation profiles into outliers without relying on external references or arbitrary genome binning. When phased long-read data are available, it also annotates haplotype-specific effects, enabling direct exploration of cis-acting rare variants. Benchmarking on simulated data confirmed robust sensitivity across realistic conditions, with sequencing depth and noise as the primary determinants of power and specificity. Using METAFORA, we conduct a systematic demonstration of methylation outlier calling at population scale jointly analyzing both PacBio and ONT long-read sequencing.

We further applied METAFORA to lrGS data from rare disease patients and family members in the UDN and GREGoR consortia. Methylation profiles were largely concordant across sequencing technologies and sites, with observed platform differences explained primarily by effective depth (a function of sequencing depth and per-read error rate). The stability of methylation across sequencing centers contrasts with other functional omics, such as RNA-seq or proteomics, which are more vulnerable to batch effects. These results highlight the relative robustness of DNA methylation as a layer for integrative analysis; this may allow analysis of single one-off samples if a technology- and tissue-matched reference is provided, lowering the barrier to apply METAFORA without having to generate a population-scale cohort. Our comparisons also show that ONT R10 chemistry substantially improves methylation detection relative to R9, with lower estimated error rates and higher concordance. R10 also outperformed PacBio HiFi in our dataset, though it is notable that our PacBio genomes were generated using an older SMTRbell prep kit 3.0 chemistry; benchmarking of newer PacBio chemistries (e.g. SPRQ) in large cohorts will be important for future evaluations.

Across the GREGoR/UDN cohort, METAFORA identified tens of thousands of outliers enriched in regulatory regions such as promoters, enhancers, and CpG islands. Haplotype-specific analyses linked many outliers to rare variants, including a deletion near TMEM97, demonstrating the power of long-read phasing to connect local methylation changes with cis-acting variation. In particular, we found CGG STR repeat expansions like in FAM193B and TOP6BL as well as CpG-rich VNTRs potentially driving allele-specific outliers.

FAM193B repeat expansion is of particular interest given prior studies implicating this locus in oculopharyngodistal myopathy (OPDM) (57,58). Rare expansions at this locus have been observed previously in population databases, in some cases accompanied by local hypermethylation (59,60). Notably, in a prior study we identified an OPDM family where affected siblings shared an approximately 196-copy repeat expansion that showed no evidence of hypermethylation and instead exhibited marked overexpression of FAM193B (42). In contrast, the substantially larger 675-copy CGG repeat identified here in an unaffected GREGoR participant was associated with strong transcriptional silencing, including a hypermethylation outlier, reduced chromatin accessibility by ATAC-seq, and underexpression by RNA-seq. This observation is consistent with reports at another OPDM-associated locus, GIPC1, where all affected individuals carried CGG repeats of approximately 73–164 copies, whereas an asymptomatic individual harbored a substantially larger ∼500-copy expansion (61). The pattern also parallels the CGG repeat at the FMR1 locus, where intermediate “premutation” repeats lead to Fragile X-associated tremor/ataxia syndrome mediated by repeat translation, while larger expansions (>200 copies) trigger promoter hypermethylation, FMR1 silencing, and Fragile X syndrome (62,63). We speculate that sufficiently large FAM193B repeat expansions may induce epigenetic silencing that protects against translation of toxic polyglycine repeat products (64), potentially rendering some very large alleles clinically benign. However, further validation in additional OPDM families and population cohorts will be required to evaluate this model.

Methylation outliers were also enriched for overlap with other functional omics outliers (chromatin accessibility, expression, protein), underscoring how they can be used to identify loci with broad regulatory disruption. Two case studies highlight the clinical potential: imprinting-associated outliers at KCNQ1OT1 and GNAS could be phased and directly linked to structural variants, providing candidate diagnoses for patients with unexplained syndromic phenotypes. Together, these examples show how methylation outlier analysis can extend genome sequencing beyond variant discovery to functional interpretation.

The main limitation of this work is a lack of a genome-wide truth set for methylation for methylation comparisons. We used intraplatform correlation and mitochondrial CpGs (which are reliably unmethylated) as proxies, but these mitochondria estimates only approximate error rates in extreme hypomethylation contexts and may not fully generalize to nuclear methylation.

Despite this, previous sequencing benchmarks using oxidative bisulfite-sequencing as a gold standard have confirmed trends we observed, including major improvements from ONT R10 over R9 (65,66), and better performance of R10 compared to PacBio HiFi (22). Our simulations also highlighted scenarios in which METAFORA is sensitive to false positives—particularly high-depth, and high-noise data—potentially explaining the excess of outliers in our PacBio cohort, though it is possible PacBio uncovered more true positives as well. Parameter tuning and improved filtering will be valuable for reducing such artifacts and streamlining curation.

Future improvements to METAFORA could include explicit modeling of haplotype-specific methylation to increase power in imprinting regions and minimize false positives driven by coverage imbalance. Future work could integrate methylation outliers into the Watershed framework to automate the prioritization of rare variants that drive epigenetic changes, thereby aiding variant interpretation (42,67,68). As population long-read sequencing continues to scale through initiatives like All of Us, methods to extend METAFORA to cohorts of tens of thousands of genomes will be needed. Such efforts will create new opportunities to map aberrant epigenetic regulation across diverse populations and connect it to rare genetic variation.

### Conclusions

In conclusion, METAFORA provides a framework for detecting and interpreting methylation outliers from long-read sequencing, enabling allelic resolution and integration with multi-omics data. By linking outliers to rare variants and downstream regulatory effects, it moves long-read methylation beyond descriptive epigenomics toward actionable insight. We envision methylation outlier analysis becoming an important bridge between genome sequencing and rare disease diagnosis, expanding the scope of clinical interpretation to regulatory and imprinting mechanisms often invisible to standard methods.

## Supplementary Methods

### Outlier simulation

Outliers were simulated across a large range of parameters following the code in “outlier_simulation_benchmark/outlier_simulation.R”. In brief, we used mean population betas across chr1 as a background. We selected a random CpG within the interior of the chromosome and flanked that site on either side by a specified number of CpGs given by the ***context_length*** parameter (default: 1,500). Into this region, we spiked in an outlier segment of length ***M*** (default: 20 CpGs), and set the expected methylation beta at these CpGs to the specified ***pop_median*** parameter, with additional variance introduced by the **noise** parameter to reflect small deviations among consecutive CpGs. This profile defines the expected “true” population methylation levels for control samples.

We then simulate a single outlier sample, by adding an outlier ***delta*** parameter to all CpGs within the designated outlier segment to give us the expected outlier methylation proportions for the region. Methylation betas for controls and the outlier sample were winsorized to the range [*1/D, (1-D)/D*], where ***D*** is the depth parameter, to prevent beta values reaching extremes of greater than one or less than zero. These vectors define the expected “true” population betas for both control samples and the outlier sample. To simulate observed betas, methylation proportion values were drawn from a beta distribution with shape parameters determined by the expected population beta and the simulated CpG depth for each of the *N-1* controls and the 1 outlier sample. To simulate CpG depths, values were drawn from a Poisson distribution parameterized by the overall mean depth parameter ***D*** across all CpGs and samples. To account for technical sources of noise in estimating observed betas, we additionally added Gaussian noise scaled by the ***noise*** parameter to the observed betas.

We simulated beta and depth matrices for all control samples and the outlier sample across a range of parameters for population median beta, outlier delta, ***N*** (number of samples), ***M*** (number of CpGs in the outlier segment), ***D*** (mean sequencing depth), and the noise parameter. We then ran METAFORA outlier calling on 10 replicates for each combination of parameters. To evaluate outlier detection performance, we calculated the overlap between outlier regions identified by METAFORA and the expected true outlier coordinates. Outlier regions with no overlap with the true outlier were classified as false positives. Outlier regions with at least 50% reciprocal overlap with the true outlier were classified as true positives.

### Methylation technology comparison

To compare genome-wide estimation of methylation proportions across different long-read sequencing platforms, we collated long-read data from the Undiagnosed Disease Network and GREGoR consortium. BAM files containing methylation marks were downloaded from UDN and GREGoR dbGaP under the following accession numbers (phs003047,phs001232). For the technology comparison section we restricted to samples sequenced from whole blood, which included 77 ONT R9 samples (68 sequenced from Stanford UDN, 9 sequenced from UW GREGoR site), 98 ONT R10 samples (8 sequenced from Stanford UDN, 15 sequenced from UW GREGoR site, 17 sequenced from UCI GREGoR site, and 58 sequenced from Broad GREGoR site), and 376 PacBio samples (286 sequenced from Stanford GREGoR site, 90 sequenced from UCI GREGoR site).

METAFORA processing (see METAFORA methods 1) were applied on each bam file according to whether it was PacBio or ONT to get genome-wide methylation beta and depth matrices for each sample. Segmentation algorithm was performed as described in METAFORA Methods 2.2, to segment genome-wide CpG methylation profiles into correlated blocks of contiguous CpGs to reduce dimensionality. These CpG blocks were used to identify and exclude 10 global outlier samples based on low mean pairwise correlations < 0.95, including 3 ONT R9 samples with low coverage (<15x) and 7 ONT R10 samples from Broad’s GREGoR site that were potentially misannotated and sequenced from a tissue other than whole blood (such as PBMCs or saliva). For each technology the mean beta across all technology samples was calculated as well as the mean effective depth, number of reads where methylation was confidently called. Segments were labeled based on mean segment beta across all samples as either hypomethylated (beta < 0.25), hypermethylated (beta > 0.75), or otherwise variably methylated. PCA was also performed on the segment beta matrix using the pcatools package in R.

For the mitochondrial error rate estimation, we first filtered long-read bams to only primary alignments aligned to chrM, removing supplementary and non-primary reads that could reflect nuclear mitochondrial DNA insertions. From these filtered bams, we calculated methylation beta and depth matrices for all samples, and then assigned the estimated error rate to be the percentage of reference methylated CpGs over all primary alignments, the main assumption being mitochondrial DNA should not have any CpG methylation. These estimated error rates were then summarized and compared across sequencing technologies.

To find differentially methylated segments between PacBio and ONT, we first excluded R9 samples to filter down to 376 PacBio samples and 91 R10 samples. We used the R package limma (lmFit) to run a linear model for each autosomal methylation segment to predict M-transformed methylation levels on an indicator variable for if the sample was sequenced on PacBio. We then labeled each segment based on the absolute difference between the median ONT beta and the median PacBio beta. Significantly deviating segments were defined as those that had an absolute technology difference of greater than 0.1 (10 percentage points difference in either technology).

### Outlier characterization and enrichment analysis

To test METAFORA on real world data, we used long-read data from UDN and GREGoR as described above as input into METAFORA. We ran METAFORA with minimum outlier size of 30 CpGs and minimum absolute outlier delta of 0.3 to focus on the largest effect deviations. We calculated burden of outliers per genome and removed 10 samples who were global outliers (>165 outliers per genome, corresponding to a scaled outlier burden z-score of 3). To functionally characterize outlier regions that were output by METAFORA, we first created a background set of non-outlier regions by sampling 100,000 segments from the genome wide population mean segmentation. These background regions were then compared to outlier segments based on their length and mean beta across all samples. To assess noncoding constraint, gnocchi non-coding constraints z-scores for 1kb bins of the genome were downloaded from https://hgdownload.soe.ucsc.edu/gbdb/hg38/gnomAD/mutConstraint/mutConstraint.bw (46). Segments were summarized as the mean gnocchi z over all overlapping bins.

To compute segment-level enrichments based on functional annotations, we compiled a list of functional annotations. These included CpGIslandsExt from UCSC table browser https://genome-euro.ucsc.edu/cgi-bin/hgTables, for CpG Islands. These islands were flanked on either side by 2Kb to define CpG shores. And additionally flanked an additional 2Kb from these shores to define CpG shelfs. ABC enhancers from all cell types were downloaded from following FTP server (https://mitra.stanford.edu/engreitz/oak/public/Nasser2021/AllPredictions.AvgHiC.ABC0.015.minus150.ForABCPaperV3.txt.gz). LiftOver was used to map these hg19 coordinates onto hg38, and then enhancers with the same target gene and type (promoter, genic, intergenic) were merged across also cell type. The Merged GRCh38 coordinate ABC enhancers can be downloaded here (https://github.com/tjense25/METAFORA/blob/main/references/ABC_enhancers.merged.GRCh38.bed.gz) (40,69). GREGoR PBMC ATAC data was joint called using ENCODE pipeline to define PBMC open chromatin, available for download here (https://github.com/tjense25/METAFORA/raw/main/references/GSS_PBMC_ATACpeaks.open_chromatin.GRCh38.bed.gz) (70). Finally, ENCODE cis conserved regulatory elements (cCRE) were downloaded from UCSC table browser https://genome.ucsc.edu/cgi-bin/hgTrackUi?db=mm10&g=encodeCcreCombined (ENCODE).

We label our outlier and background segments based on if it overlaps each annotation (plyranges overlapsAny function). To calculate enrichments, we then apply fisher’s exact test to get odds ratios, confidence intervals, and p-values.

### Multi-omics enrichments

To calculate enrichments across multiple omics layers, we use additional multi-omics data available from UDN and GREGoR as described below. We computed these enrichments at the gene level to make it consistent across the different layers of regulation. To define methylation outlier genes, we took gene coordinates from GENCODE v32, subset to protein coding genes, slopped them with an additional 10Kb on either side, and then intersected with methylation outlier coordinates from METAFORA output for the 494 blood tissue samples. The gene in each sample was labeled binary as containing an outlier nearby or not. To compute pairwise multi-omics enrichments we then subset to samples present in both assays, and subset to genes that had at least one outlier sample in both assays. Fisher’s exact test was then used on gene counts across the contingency table of outlier presence across each of the two omes to get enrichment odds ratio, confidence interval, and p-value.

#### Rare SVs

We use the same 551 UDN and GREGoR input genomes to METAFORA as input to call structural variants. Sniffles2 (71) was called on each long-read sample bam using recommended default parameters. Resulting SV vcf files were merged with jasmine as previously described (42,72). From the joint SV file, allele frequencies were estimated from observed allele counts and then variants filtered to those with minor allele frequency < 1%. Breakends (SVTYPE = “BND”) and SV calls smaller than 50bp were also filtered out. As quality control, we removed 6 samples which contained an excess number of breakends or SVs (defined as samples with > 5 sd above mean count per genome for any SV type). We intersected rare SV coordinates with slopped gene coordinates to define rare SV genes. In the SV-methylome enrichment, we calculated enrichment based on 484 shared samples across 4,575 shared genes.

#### ATAC-seq Chromatin Accessibility Outliers

To call chromatin accessibility outliers, we followed methods recommended by EpiOut (8). First, we started with ATAC bam files for 184 GREGoR participants processed with the ENCODE pipeline (70). We downsampled and then merged reads from all bams into a single monolithic bam file. This file was then run through MACS2 peak calling to define a joint peak set. This joint peak set was then intersected with sample-level bams to get a count matrix across all samples and peaks. This count matrix was then used as input to EpiOut with default parameters to yield chromatin accessibility outliers. EpiOut reports log2 fold change of each ATAC peak defined as log2 ratio of observed counts divided by an autoencoder-corrected predicted expected count.

EpiOut also reports a p-value per peak that this fold change deviance is significantly different from the null hypothesis. We define outliers as peaks where the multiple-testing corrected p-value, *padj*, is less than alpha threshold 0.05. As quality control, we removed 2 samples that displayed a higher than expected number of outliers per genome (defined as > 3sd above mean expected burden of outliers). We intersected outlier peak coordinates with slopped gene coordinates to define ATAC outlier genes. In the ATAC-methylome enrichment, we calculated enrichment based on 155 shared samples across 1,058 shared protein-coding genes.

#### RNA-seq Expression Outliers

To call expression outliers we followed the same procedure as previously described for RNA-seq data (42,73). We started with 455 GREGoR and UDN RNA bam files sequenced from whole blood aligned to hg38 with STAR. Gene counts were quantified with RNASeQC, corrected for library size and composition, and then log-normalized. PCA was used to estimate hidden factors, and log-scaled counts were then residualized on these top 50 PCs, along with sex and sequencing batch indicator variables. Finally, residuals per gene were centered and scaled to get expression z-scores. Expression outliers were defined as any gene with absolute z-score > 2. As quality control, we removed 9 samples that were global outliers, containing an excessive burden of outliers (>3sd above mean expected burden of outliers per genome). In the RNA-methylome enrichment, we calculated enrichment based on 108 shared samples across 3,832 shared protein-coding genes.

For isoform-level expression z-scores in the GNAS example, a similar process was performed but using RSEM to quantify genes based on STAR transcriptome-aligned bams. Specific GNAS transcript *ENST00000485673* was chosen for analysis as its TSS overlaps imprinting and outlier region and it contained unique exonic sequence to enable isoform quantification.

#### OLink Proteomics Outliers

To call proteomics outliers, we used plasma protein abundance data from Olink HT assay available for 172 GREGoR participants. To normalize data per protein, we started with PCNormalizedNPX values generated through Olink’s workflow. These NPX values are normalized intensities to plate controls that are log-scaled and residualized on PCs to correct for additional technical factors. GREGoR sample proteomics were generated from 3 separate plates, so to further correct potential batch effects, we performed additional PC hidden factor residualization. PCA was performed on the matrix of PCNormalizedNPX values across all samples, and top 20 PCs along with indicator values for the different plateID were corrected with a linear model. Residuals were then scaled and centered to obtain protein z-scores. We defined protein outliers as a protein with absolute z-score > 2. To assign proteins to genes, UniProt IDs provided by assay were mapped to ENSEMBL gene ids and hgnc symbols using biomart. We did not observe any major global outliers in the proteomics data in terms of outlier burden per genome. In the Protein-methylome enrichment, we calculated enrichment based on 120 shared samples across 1,361 shared protein-coding genes.

## Declarations

### Ethics approval and consent to participate

This study was approved by the Stanford University Institutional Review Board (protocol 60837) and the National Human Genome Research Institute Institutional Review Board (protocol 15-HG-0130). All participants or their legal guardians provided written informed consent for participation in genomic research and data sharing.

### Consent for publication

As part of the above IRB protocols, all participants or their legal guardians provided informed consent for publication of de-identified genomic and clinical data.

### Availability of data and code

METAFORA source code, workflows, and analysis scripts are available at the METAFORA github, along with instructions for its implementation (https://github.com/tjense25/METAFORA). Long-read sequencing data and multi-omics for the 551 samples used in this paper are available through dbGaP at the following accession numbers. All GREGoR samples available at phs003047, and all UDN samples available at phs001232.

### Authors’ contributions

TDJ conceived the study, developed the METAFORA codebase, performed analyses, interpreted results, and drafted the manuscript. RK and JN assisted in data processing, analysis, and interpretation. DEB and CMR assisted with clinical interpretation. JAB and MTW contributed clinical interpretation and oversight of rare disease analyses. EAA and SBM supervised the study and revised the manuscript. All authors reviewed and approved the final manuscript.

## Acknowledgments

We extend thanks to all research participants in the UDN and GREGoR consortia. This work required computing resources from the Stanford Genetics Bioinformatics Service Center (supported by NIH Instrumentation Grant S10 OD025082). We kindly thank Rachel Ungar for helping process RNA-seq and obtaining expression z-scores and Jeren Olsen for helping process ATAC-seq data and running EpiOut.

---

**Supplemental Figure 1:**
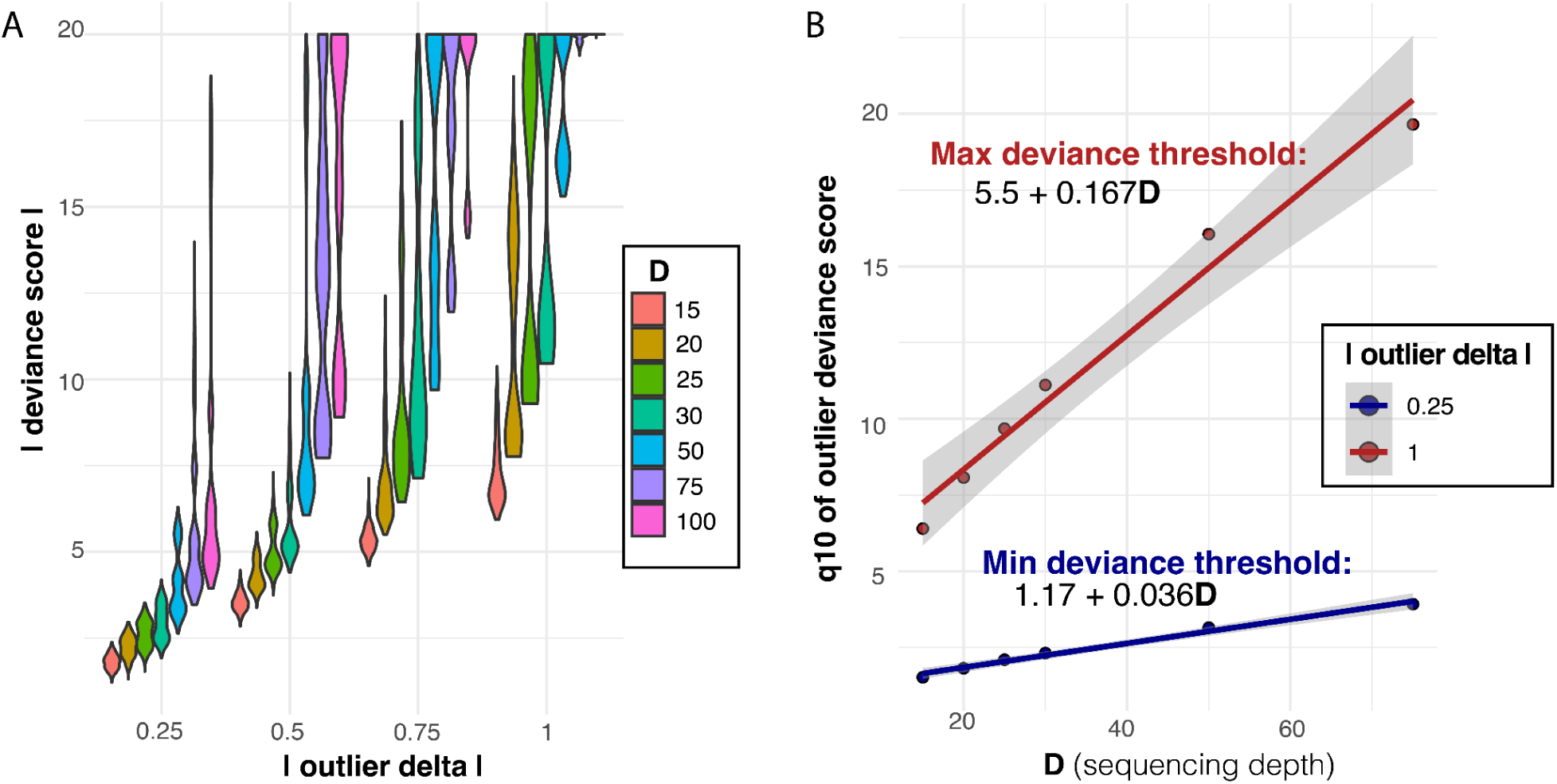
learning thresholds for deviance score from simulated outliers. A Distribution of deviance scores at various outlier deltas for simulated outliers across a range of depth thresholds. As depth increases so do absolute deviance scores. B Linear relationship between depth and the 10th quantile of observed deviance score from simulated outliers. Q10 at an absolute delta of 0.25 is set as a minimum deviance score threshold to ensure 90% power at detecting outliers of magnitude 0.25. Q10 at an absolute delta of 1.0 is set as maximum deviance score to avoid extreme inflation and infinite values of deviance score. Slope and intercept for these depth-dependent thresholds are annotated from fitting linear models.

**Supplemental Figure 2.**
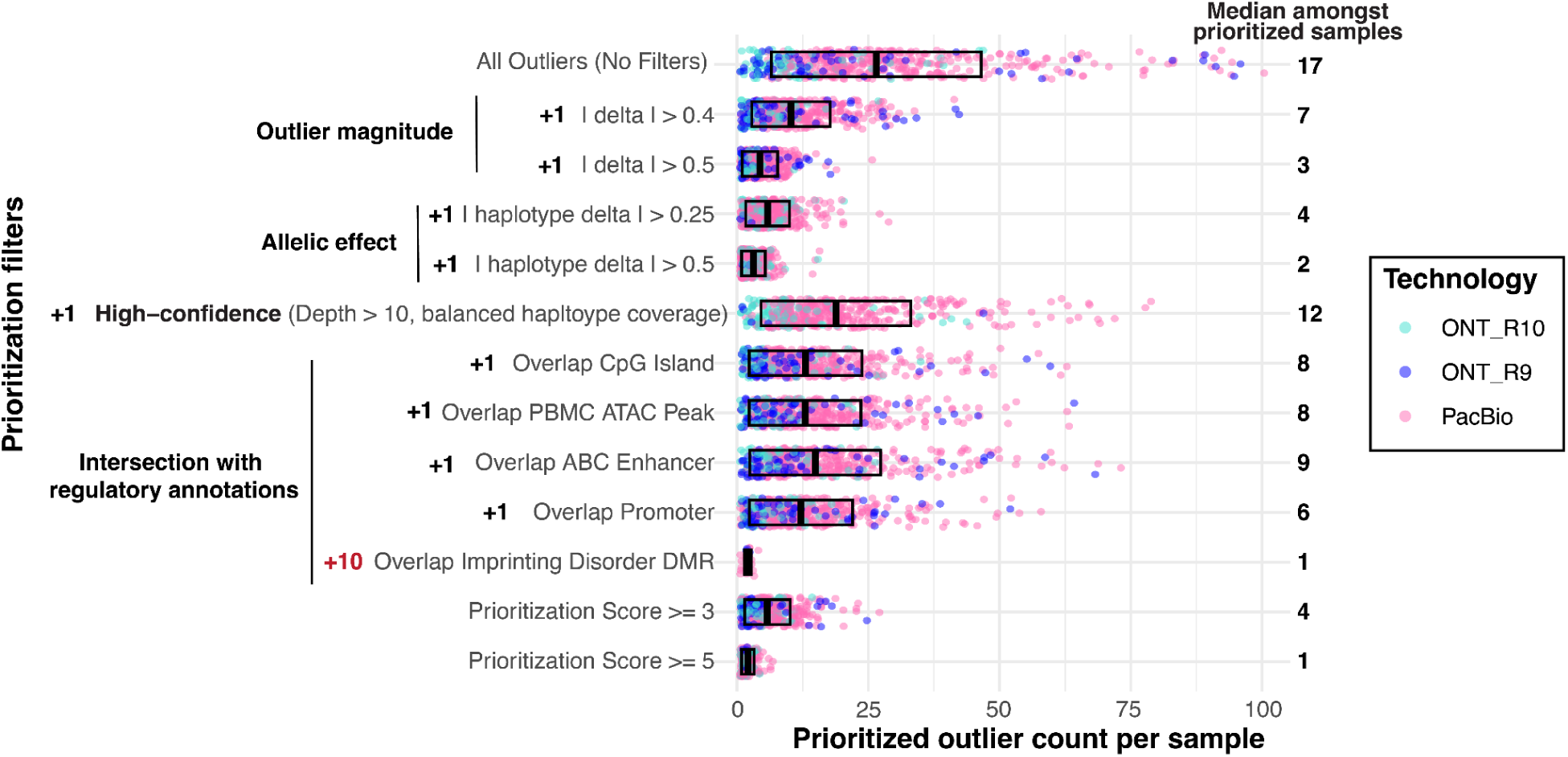
Outlier prioritization scores. Outlier prioritization from METAFORA applied to GREGoR U08 and UDN long-read data. X-axis displays the number of outliers per sample that meet the corresponding prioritization filter on Y-axis, colored by sequencing technology. Outliers gain the specified number of points for meeting filter criteria. Median outlier count per sample annotated for each filter, amongst samples with at least one outlier meeting filter. At the end of scoring, samples had a median of 4 outliers with a prioritization score of at least 3 and 1 outlier with prioritization score of at least 5.

**Supplementary Figure 3.**
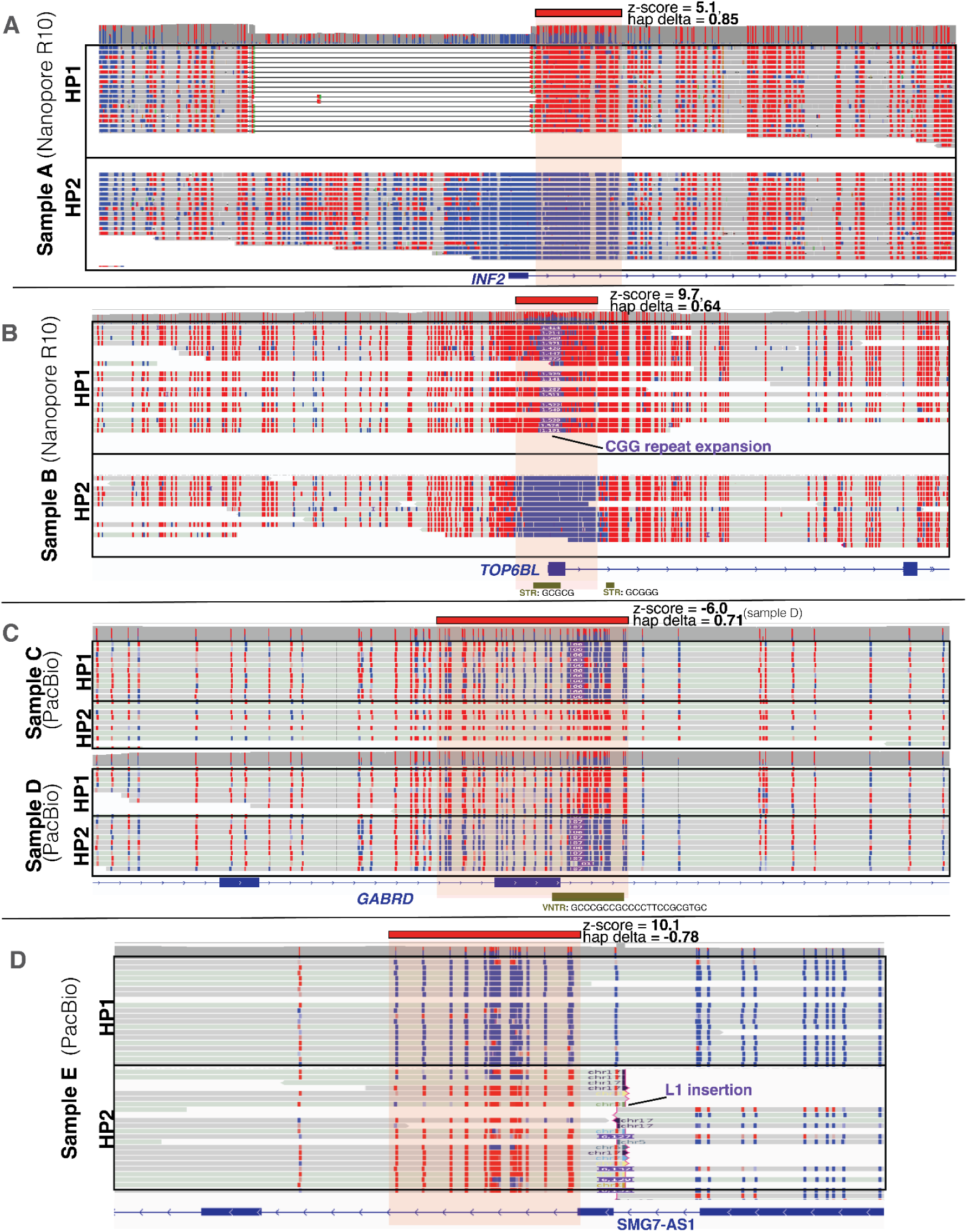
IGV screenshots displaying high haplotype delta outliers near rare structural variants. A 1.7Kb deletion of the promoter region of *INF2* observed upstream of a hypermethylation outlier in a nanopore R10 sample. B 1.5Kb CGG repeat expansion insertion in promoter/5’UTR of *TOP6BL* gene phased together with a hypermethylation outlier in a Nanopore R10 sample. C Rare insertions in a VNTR region intronic to the *GABRD* identified in multiple samples. Sample C sequenced from PacBio has 3 additional copies of 22bp repeat motif (z-score=-4.52), and is associated with hypomethylation of that haplotype. Sample D also from PacBio has additional 4 copies and even stronger hypomethylation (z-score=-5.99). D A rare 6Kb L1 retrotransposon insertion identified and phased together with an upstream hypermethylation outlier of an *SMG7-AS1* intron in a PacBio sample. In all IGV screenshots gene models are displayed below along with adotto catalog of tandem repeat loci and their annotated motif.

